# Associations of ABO and Rhesus D Blood Groups with Phenome-Wide Disease Incidence: A 41-year Retrospective Cohort Study of 482,914 Patients

**DOI:** 10.1101/2022.09.23.22280292

**Authors:** Peter Bruun-Rasmussen, Morten Hanefeld Dziegiel, Karina Banasik, Pär Ingemar Johansson, Søren Brunak

## Abstract

**Background:** The natural selection responsible for the observed blood group frequency differences between populations remains debatable. The ABO system has been associated with several diseases and recently also with susceptibility to COVID-19 infection. Associative studies of the RhD system and diseases are sparser. A large disease-wide risk analysis may further elucidate the relationship between the ABO/RhD blood groups and disease incidence.

**Methods:** We performed a systematic log-linear quasi-Poisson regression analysis of the ABO/RhD blood groups across 1,312 phecode diagnoses. Unlike prior studies, we determined the uniqueness of each individual ABO blood group as opposed to using blood group O as the reference. Moreover, we used up to 41-years of nationwide Danish follow-up data, and a disease categorization scheme specifically developed for diagnosis-wide analysis. Further, we determined associations between the ABO/RhD blood groups and the age at the first diagnosis. Estimates were adjusted for multiple testing.

**Results:** The retrospective cohort included 482,914 Danish patients (60.4% females). The incidence rate ratios (IRRs) of 101 phecodes were found statistically significant between the ABO blood groups, while the IRRs of 28 phecodes were found statistically significant for the RhD blood group. The associations included cancers and musculoskeletal-, genitourinary-, endocrinal-, infectious-, cardiovascular-, and gastrointestinal diseases.

**Conclusions:** We found associations of disease-wide susceptibility differences between the blood groups of the ABO and RhD systems, including cancer of the tongue, monocytic leukemia, cervical cancer, osteoarthrosis, asthma, and HIV- and hepatitis B infection. This may indicate that the ABO/RhD blood groups are the results of a selective pressure driven partly by robustness to disease. We found marginal evidence of associations between the blood groups and disease onset.

**Funding:** Novo Nordisk Foundation and the Innovation Fund Denmark

## Introduction

Still 100 years after the discovery of the ABO and Rhesus D (RhD) blood group systems, the selective forces responsible for the observed blood group population differences remain elusive.^1^ The pathophysiological mechanisms behind the observed relationship between blood groups and diseases are not well understood either. The ABO system has been associated with susceptibility to multiple diseases, including gastrointestinal- and cardiovascular diseases and pancreatic-, gastric-, and ovarian cancers.^2,3,4,5,6,7,8^ The ABO system has also been associated with the susceptibility, progression, and severity of COVID-19.^9^ In contrast, apart from hemolytic disease of the newborn, reported associations between the RhD blood group and disease development are sparser.^1^

Specifically, higher levels of factor VIII (FVIII) and von Willebrand factor (vWF) observed in individuals with a non-O blood group have been suggested to affect the development of cardiovascular disease.^10, 11^ Additionally, blood group-related antigens have been suggested to be involved in the adhesion of trophoblast, inflammatory cells, and metastatic tumor cells to the endothelial cells of the vasculature.^12^ The endothelial cells of the vasculature have also been suggested to contribute to the initiation and propagation of severe clinical manifestations of COVID-19.^13^

Recently, an associative disease-wide risk analysis of the ABO and RhD blood groups was conducted in a large Swedish cohort.^7^ The study generated further support for previous findings and suggested new associations. Here, we further uncover the relationship between the ABO and RhD blood groups and disease susceptibility using a Danish cohort of 482,914 patients. In contrast to previous studies, we use up to 41-years of follow-up data, and a disease categorization scheme specifically developed for diagnosis-wide analysis called phecodes.^14^ Further, we determine the uniqueness of each individual ABO blood group as opposed to using blood group O as the reference.

We estimate incidence rate ratios of 1,312 phecodes (diagnoses) for the ABO and RhD blood groups. Further, we determine associations between the ABO/RhD blood groups and the age at the first diagnosis to better disclose the temporal life course element of disease development.

## Methods

### Study design

This retrospective cohort study was based on the integration of the Danish National Patient Registry (DNPR) and data on ABO/RhD blood groups of hospitalized patients. We included Danish patients who had their ABO/RhD blood group determined in the Capital Region or Region Zealand (covering ∼45% of the Danish population^15^), between January 1, 2006, and April 10, 2018. The DNPR provided the International Classification of Diseases 8^th^ and 10^th^ revision (ICD-8 and ICD-10) diagnosis codes, dates of diagnosis, date of birth, and sex of patients, with records dating back to 1977.

Similar to a case-control study, the patients were included retrospectively. Here, selection into the study was based on an in-hospital ABO/RhD blood group determination. That is, the person-time and the entire disease history back to 1977 of patients hospitalized between 2006-2018 with known ABO/RhD blood groups were included retrospectively.

We defined diseased and non-diseased individuals using the phecode mapping from ICD-10 diagnosis codes.^14^ Before categorizing the assigned ICD diagnosis codes into phecodes, the ICD-8 codes were converted to ICD-10 codes.^16^ Further, referral diagnoses were excluded. Pregnancy- and perinatal diagnosis (ICD-10 chapters 15-16) assigned before or after age 10 were excluded, or, when possible, rightly assigned to the mother or newborn, respectively. The disease categories of injuries, poisonings, and symptoms were deemed unlikely to be associated with the blood groups and excluded from the analyses (phecode categories: “injuries and poisonings”, “symptoms” and phecodes above 999). To ensure sufficient power, only phecodes with at least 100 cases in the study sample were included. The patients were followed from the entry in the DNPR to the date of death, emigration, the first event of the studied phecode, or end study period (April 10, 2018), whichever came first. Thus, follow-up was up to 41 years. The patients were allowed to contribute events and time at risk to multiple phecode analysis.

### Diagnosis-wide incidence rate ratios

We used a log-linear quasi-Poisson regression model to estimate incidence rate ratios (IRRs) of each phecode among individuals with blood groups A, B, AB, and O relative to the other blood groups, respectively (e.g. A vs. B, AB, and O).^17, 18^ Further, we compared individuals with positive RhD type relative to negative RhD type. The analyses of diseases developed by both males and females were adjusted for sex, while analyses of sex-restricted diseases (e.g. cervical cancer) only included a subgroup of individuals of the restricted sex. Sex-restricted diseases were pre-defined by the phecode terminology. Sex was considered a confounder as prior studies have found sex differences in the incidence rates of multiple diseases.^19^ Further, we adjusted for the year of birth and attained age, both modeled using restricted cubic splines with five knots. Attained age was split into 1-year intervals and treated as a time-dependent covariate, thus allowing individuals to move between categories with time. Herewith, age was used as the underlying time scale. Further, an interaction between attained age and sex was modeled for non-sex-restricted analyses. Patients were excluded from the analysis if they were assigned the phecode under study at the start of the DNPR. For analysis of congenital phecodes (e.g., sickle cell disease), prevalence ratios were estimated instead of IRRs by using the cohort size as the offset (see Supplementary Table 1 for a list of the congenital phecodes). The analyses of ABO blood groups were adjusted for RhD type, and the RhD-analyses were adjusted for the ABO blood group. Adjustment for the birth year was done to control for societal changes and was used instead of the calendar period of diagnosis. The robust quasi-Poisson variance formula was used to control for over-dispersion.^18^

### Age of first hospital diagnosis

We estimated differences in age of first phecode of individuals with blood group A, B, AB, and O relative to any other blood group, respectively. Similar analyses were done for RhD positive individuals relative to RhD negative individuals. We used a linear regression model adjusted for sex and birth year (as a restricted cubic spline with five knots). Analysis of sex-restricted phecodes was not adjusted for sex. Individuals who were assigned the studied phecode at the start date of the DNPR were excluded as the age of diagnosis was uncertain. Further, congenital- and pregnancy related phecodes were not included.

Statistical analyses were performed in R (version 3.6.2) using the survival and rms package. P-values were two-sided. P-values and confidence intervals were adjusted for multiple testing by the false discovery rate (FDR) approach, accounting for the number of performed tests (5 blood groups times 1,312 phecodes).^20^ FDR adjusted p-values < 0.05 were deemed statistically significant. The analysis pipeline was made in python (anaconda3/5.3.0) using snakemake for reproducibility.^21^ The analyses code is available through www.github.com/peterbruun/blood_type_study. The manuscript complies with the STROBE reporting guidelines.

### Data access disclosure

Anonymized patient data was used in this study. Due to national and EU regulations, the data cannot be shared with the wider research community. However, data can be accessed upon relevant application to the Danish authorities. The Danish Patient Safety Authority and the Danish Health Data Authority have permitted the use of the data in this study; whilst currently, the appropriate authority for journal data use in research is the regional committee (“Regionsråd”). The statistical summary data used to create the tables and graphs are available as Supplementary Data 1 and Supplementary Data 2. The analysis code is publicly available through www.github.com/peterbruun/blood_type_study.

## Results

In total, 482,914 patients (60.4% females) were included and 1,312 phecodes (diagnosis codes) were examined (Figure 1, and Table 1). The median follow-up time for all phecode analyses was 17,555,322 person-years (Q1-Q3: 17,324,597 – 17,615,142). The cohort held a wide age distribution of patients born from 1901 to 2015 (Table 1, and Supplement Figure 1). The ABO/RhD blood group distribution of the patients was similar to that of a previously summarized reference population of 2.2 million Danes (Table 1).^22, 23^

**Figure 1.**
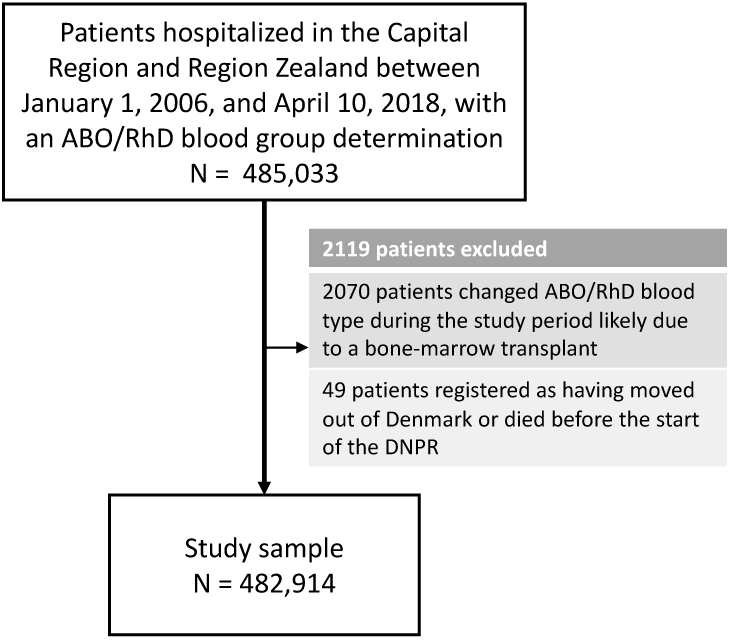
Selection of Patients for the 41-Year Retrospective Cohort Study on ABO/RhD Blood Groups and Associations with Disease Incidence in 482,914 Danish Patients.

**Table 1.** Characteristics of Patients in the 41-Year Retrospective Cohort Study on ABO/RhD Blood Groups and Associations with Disease Incidence in 482,914 Danish Patients.

| **N = 482,914** |  |  |
| --- | --- | --- |
| ABO, n (%) | O | 197,634 (40.9) |
|  | A | 206,110 (42.7) |
|  | AB | 22,111 (4.6) |
|  | B | 57,059 (11.8) |
| RhD, n (%) | Negative | 74,150 (15.4) |
|  | Positive | 408,764 (84.6) |
| Sex, n (%) | K | 291,649 (60.4) |
|  | M | 191,265 (39.6) |
| Birth year, median<br>[Q1,Q3] |  | 1963 [1945,1982] |
| Age at entry, median<br>[Q1,Q3] |  | 13 [0,32] |
| Follow-up time, median<br>[Q1,Q3] |  | 40.8 [33.4,41.3] |

### Incidence rate ratios

After adjustment for multiple testing, we found the incidence rate ratios (IRRs) of 101 and 28 phecodes (116 unique) to be statistically significant for the ABO and RhD blood groups, respectively. The statistically significant IRRs are given with 95% confidence intervals in Table 2. The estimates of all examined phecodes are given in Supplement Table 2. Further, Manhattan plots of the p-values and disease categories are presented in Figure 2.

**Table 2:**
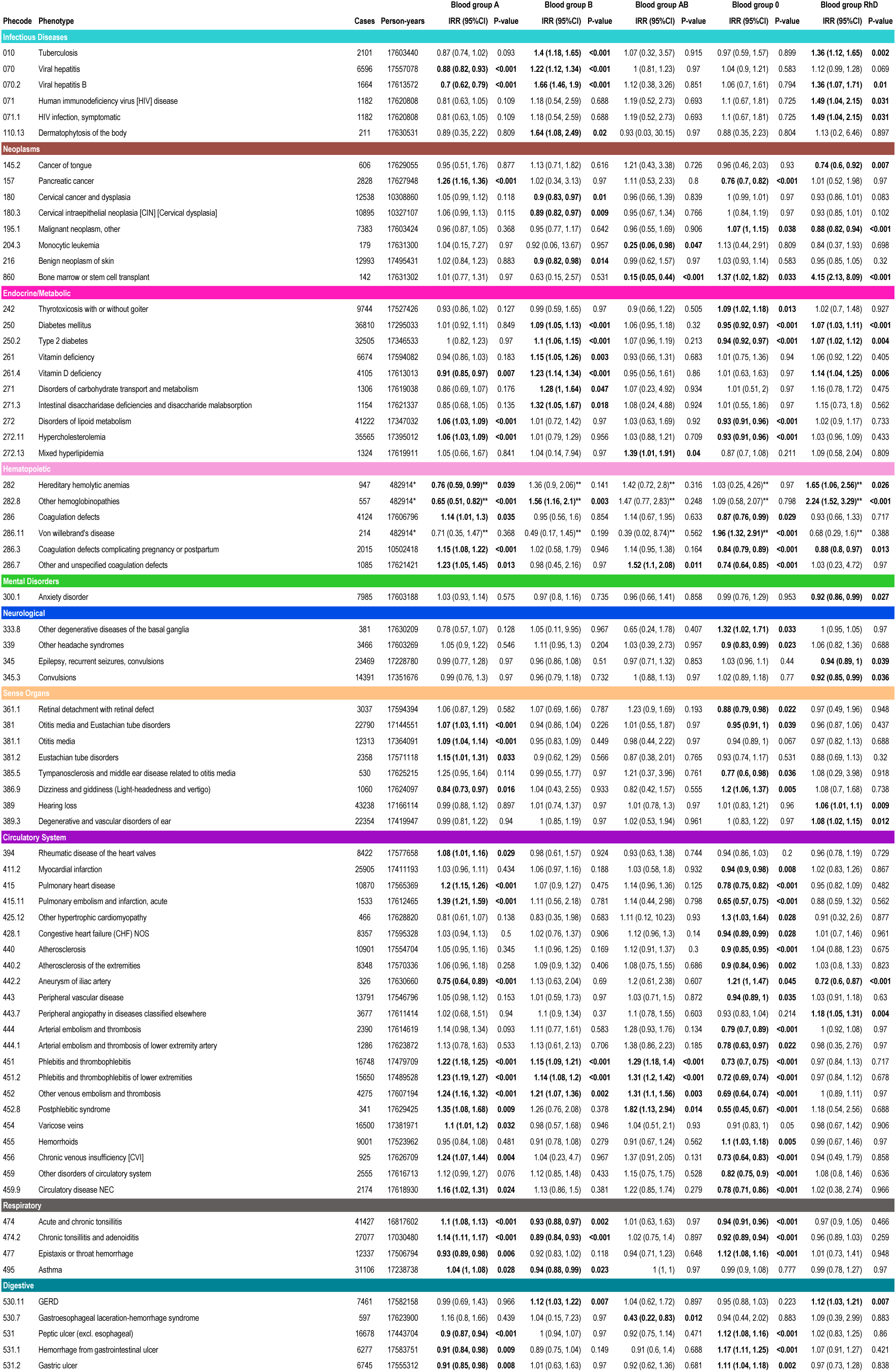
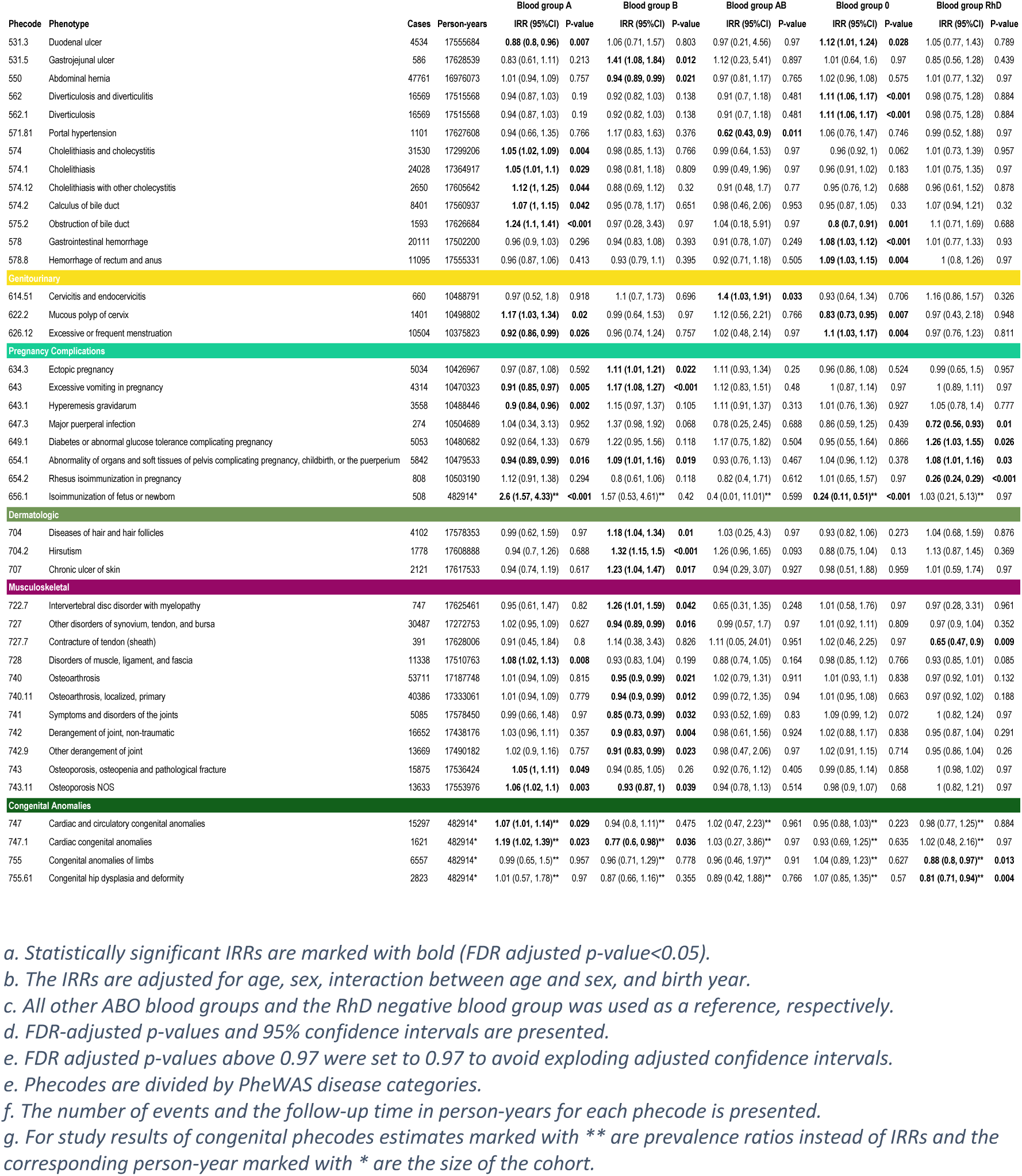
Statistically significant incidence rate ratios for each individual blood group A, B, AB, and O relative to any other blood group (e.g. A vs. B, AB, and O combined). Further, also for RhD positive blood group relative to the RhD negative blood group.

**Figure 2:**
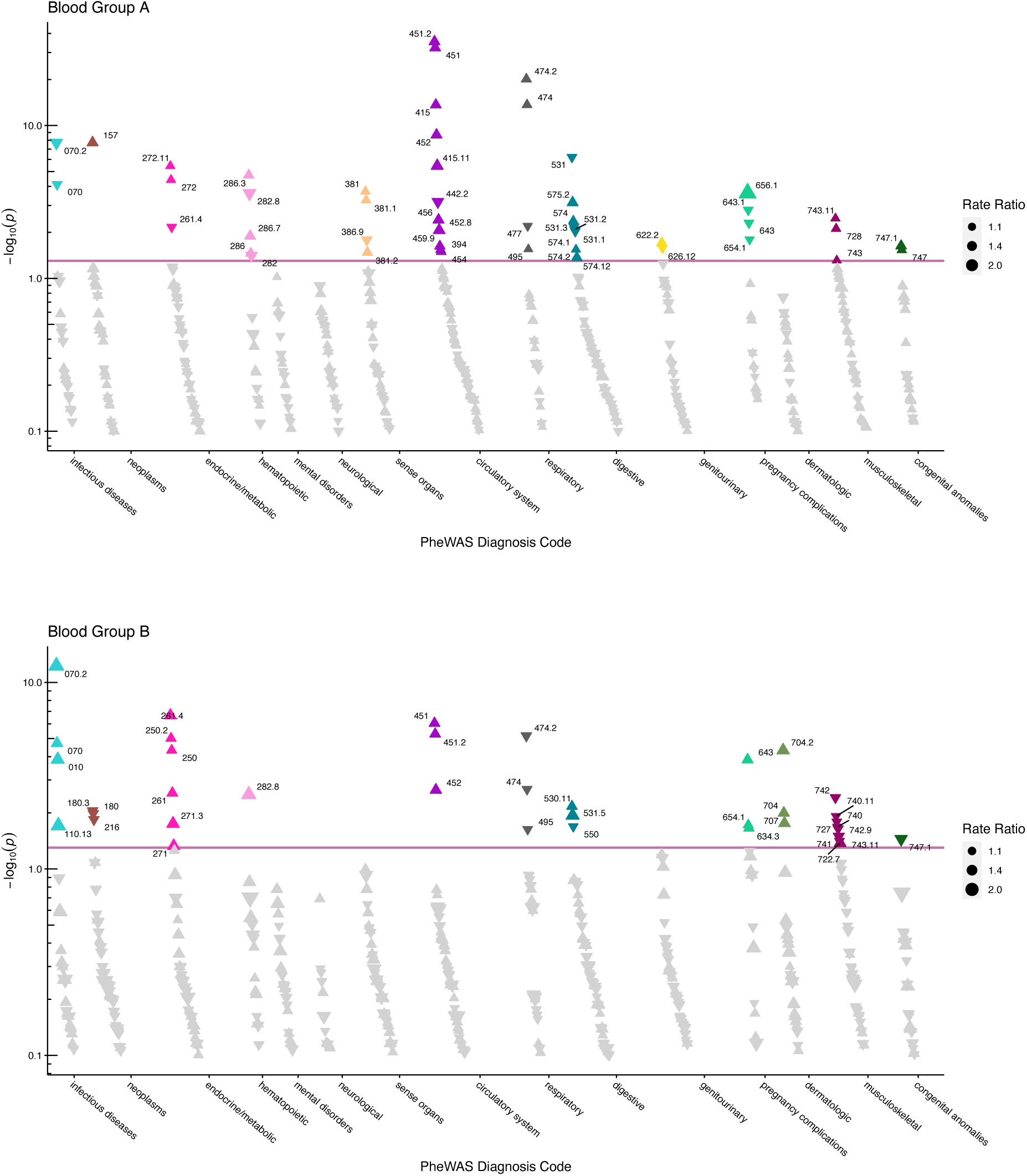
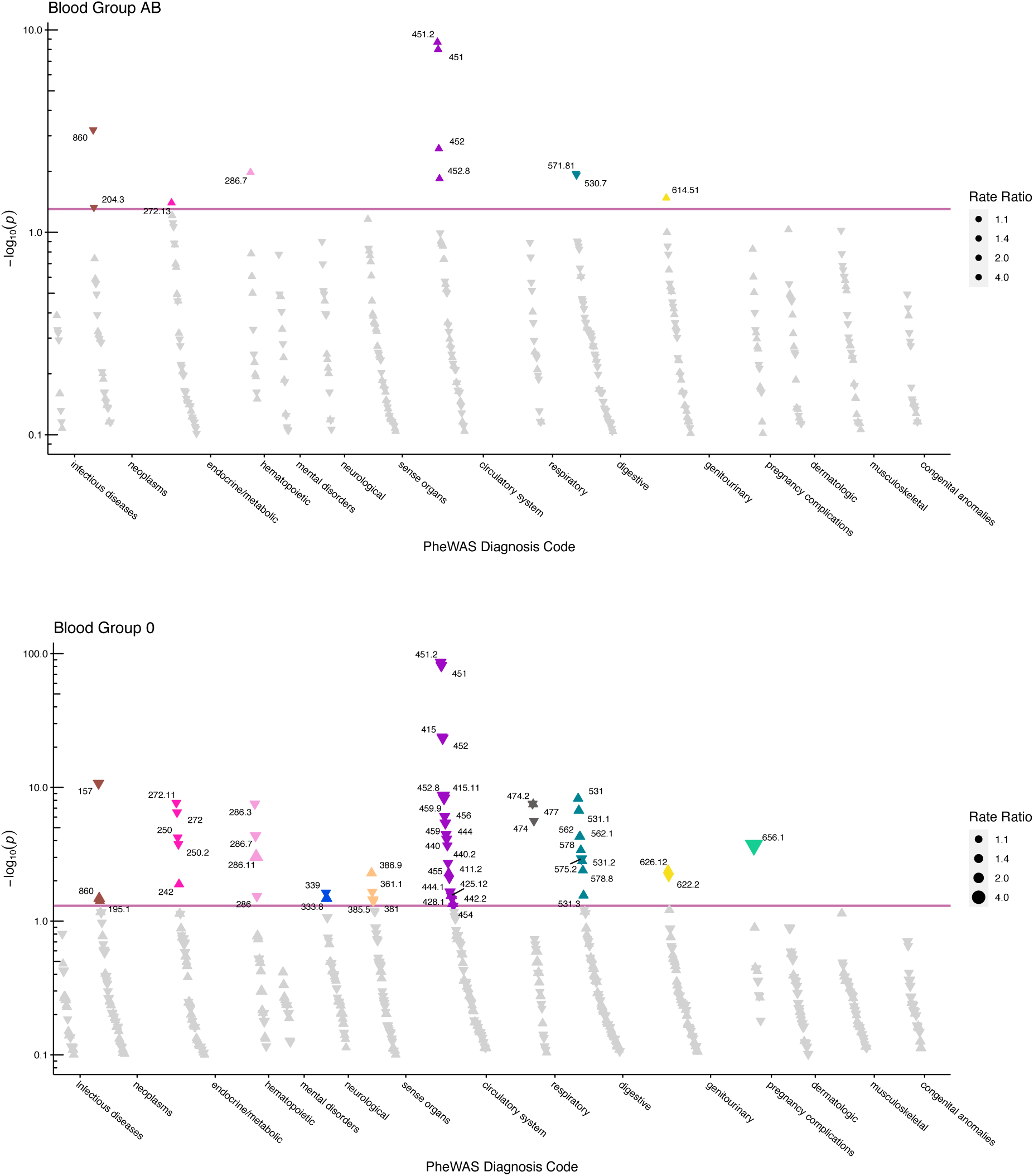
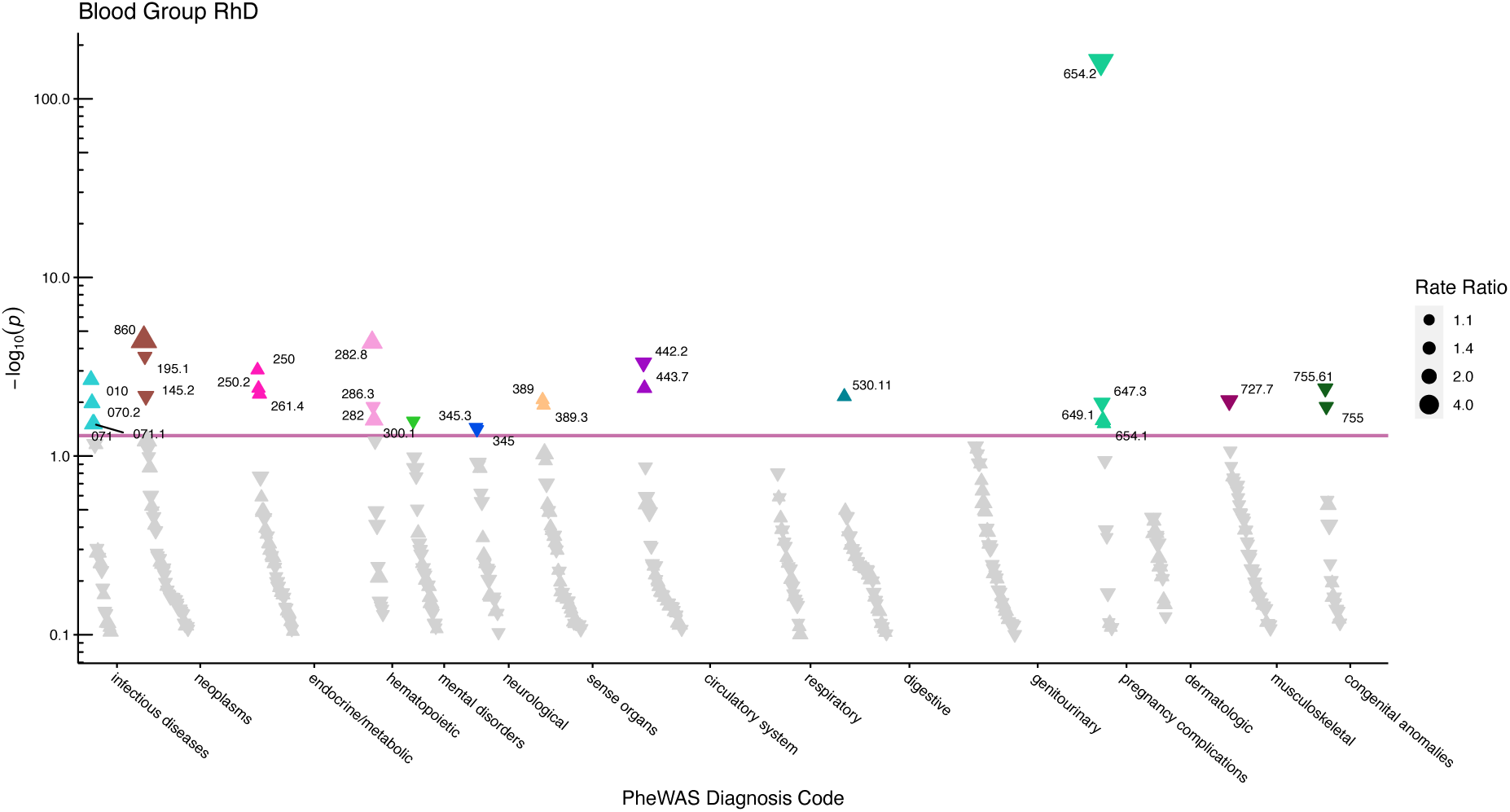
Manhattan plots for the ABO and RhD blood groups with phecodes included by category. The vertical axis shows the -log10 transformed FDR adjusted p-values on a log10-scale. The horizontal axis shows the phecodes by category. The red line indicates the statistically significant level of <0.05 for FDR adjusted p-values. Associations with p-value > 0.8 are not displayed. Coloured and annotated associations were deemed statistically significant. The direction of the triangles indicates positive or inverse associations (upward: IRR>1, downward: IRR<1). The size of the triangles indicates the size of the incidence rate ratio. A) Manhattan plot for blood group A. B) Manhattan plot for blood group B. C) Manhattan plot for blood group AB. D) Manhattan plot for blood group O. E) Manhattan plot for the Rhesus D blood group.

The number of statistically significant IRRs for A, B, AB, O, and RhD were 50, 38, 11, 53, and 28, respectively. For 13 phecodes, an association was found for both the ABO blood group and the RhD blood group. The ABO blood groups were found positively associated with 75 phecodes and inversely associated with 67 phecodes. The RhD-positive blood group was found to have 16 positive- and 12 inverse associations.

### Disease onset

We found the B blood group to be associated with a later onset of viral infection. Further, blood group O was associated with a later onset of phlebitis and thrombophlebitis (Table 3 and Supplement Table 3). The RhD positive group was associated with a later onset of acute and chronic tonsilitis diagnosis.

**Table 3:** Statistically significant associations between the ABO/RhD blood groups and the age of the first diagnosis.

| Phecode | Phenotype | N | Blood group A |  |  | Blood group B |  |  | Blood group AB |  |  | Blood group O |  |  | Blood group RhD |  |
| --- | --- | --- | --- | --- | --- | --- | --- | --- | --- | --- | --- | --- | --- | --- | --- | --- |
|  |  |  | Estimate (95%CI) | P-value |  | Estimate (95%CI) | P-value |  | Estimate (95%CI) | P-value |  | Estimate (95%CI) | P-value |  | Estimate (95%CI) | P-value |
| 079 | Viral infection | 25075 | -0.26 (-3.06, 2.54) | 0.864 |  | **0.92 (0.13, 1.71)** | **0.022** |  | 0.53 (-6.18, 7.23) | 0.887 |  | -0.23 (-3.22, 2.75) | 0.887 |  | 0.27 (-5.22, 5.75) | 0.93 |
| 451 | Phlebitis and thrombophlebitis | 16748 | **-0.58 (-1.02, -0.13)** | **0.011** |  | -0.24 (-5.71, 5.22) | 0.936 |  | -0.6 (-5.87, 4.66) | 0.833 |  | **0.91 (0.57, 1.25)** | **<0.001** |  | 0.01 (-0.33, 0.34) | 0.97 |
| 451.2 | Phlebitis and thrombophlebitis of lower extremities | 15650 | -0.53 (-1.08, 0.01) | 0.055 |  | -0.27 (-5.85, 5.31) | 0.93 |  | -0.7 (-6.35, 4.95) | 0.82 |  | **0.9 (0.55, 1.25)** | **<0.001** |  | 0.08 (-3.89, 4.06) | 0.97 |
| 474 | Acute and chronic tonsillitis | 41428 | -0.29 (-1.45, 0.87) | 0.634 |  | 0.42 (-1.97, 2.8) | 0.744 |  | 0.38 (-5.71, 6.48) | 0.909 |  | 0.05 (-2.15, 2.24) | 0.97 |  | **0.67 (0.15, 1.19)** | **0.011** |
| 474.1 | Acute tonsillitis | 18162 | 0.1 (-4.42, 4.61) | 0.97 |  | 0.41 (-7.17, 7.98) | 0.923 |  | 0.75 (-7.23, 8.74) | 0.864 |  | -0.42 (-3.76, 2.93) | 0.82 |  | **1.34 (0.64, 2.04)** | **<0.001** |

a. Statistically significant effect estimates are marked with bold (FDR adjusted p-value<0.05).
b. FDR adjusted p-values and 95% confidence intervals are presented.
c. FDR adjusted p-values above 0.97 were set to 0.97 to enable estimation of adjusted confidence intervals.
d. Estimates represent increases or decreases in years of age of first diagnosis.

## Discussion

We found the ABO/RhD blood groups to be associated with a wide spectrum of diseases including cancers and musculoskeletal-, genitourinary-, endocrinal-, infectious-, cardiovascular-, and gastrointestinal diseases. Novel associations of the ABO blood groups included monocytic leukemia, tonsilitis, renal dialysis, diseases of the female reproductive system, and osteoarthrosis. Novel associations of the RhD blood group included cancer of the tongue, malignant neoplasm (other), tuberculosis-, HIV-, hepatitis B infection, type 2 diabetes, hereditary hemolytic anemias, major puerperal infection, anxiety disorders, and contracture of tendon.

The blood groups may reflect their corresponding genetic markers; thus, our findings may indicate a linkage between disease and the ABO locus on chromosome 9 and the RH locus on chromosome 1, respectively. Alternatively, the associations may indicate that the blood groups are involved in disease mechanisms at the molecular level mediated either through the blood group antigens or by the blood group reactive antibodies. However, our findings have a compromised causal interpretation given the retrospective inclusion of individuals (and person-time) after an in-hospital blood group test.

Our results support several previously observed associations including positive associations between the non-O blood groups and prothrombotic diseases of the circulatory system (phecodes: 411.1 – 459.9), associations with gastroduodenal ulcers, associations of blood group O and lower risk of type 2 diabetes, and positive association between blood group B and tuberculosis.^2, 6, 7, 24, 25^ Further, our results support findings associating non-O blood groups with increased risk of pancreatic cancer.^3^ The role of the ABO blood group in HIV susceptibility remains controversial; we only observed a positive association for the RhD-positive blood group.^26^

We found blood group B to be positively associated with ‘ectopic pregnancy’, ‘excessive vomiting in pregnancy, and ‘abnormality of organs and soft tissues of pelvis complicating pregnancy’ indicating that blood group B mothers may be more likely to experience pregnancy complications. Further, we found positive associations of blood group A with both ‘mucous polyp of cervix’, and blood group AB with ‘cervicitis and endocervicitis’. Taken together these findings may indicate that the ABO blood groups are associated with diseases of the female reproductive system. However, the study design does not allow for any causal interpretation.

Only a few statistically significant associations were found for the analyses of the age of the first diagnosis; thus, indicating that the blood group’s involvement in disease onset may be marginal.

A strength of our approach is that we utilized the phecode disease classification scheme that is specifically developed for disease-wide risk analyses.^27^ The phecode mapping scheme combines ICD-10 codes that clinical domain experts have deemed to cover the same disease.

For example, respiratory tuberculosis (A16), tuberculosis of nervous system (A17), and miliary tuberculosis (A19), are combined into the phecode tuberculosis (phecode 10). Phecodes may therefore provide increased power and precision compared with using ICD-10 categories.^28^ Further, contrary to previous studies, we compared each blood group to all other blood groups, instead of determining effect estimates relative to blood group O. Thus, here we better capture the uniqueness of each individual ABO blood group.

### Limitations

Our study has some important limitations, firstly, the retrospective inclusion of patients and person-time may have introduced an immortal time bias from deaths before enrollment (in-hospital ABO/RhD blood group test).^29^ The findings are therefore conditioned on patients surviving until the enrollment period. This implies, e.g., that if a specific blood group causes a higher incidence of a deadly disease, then patients with such blood group are more likely to have died before enrollment, and therefore fewer individuals having both that blood group and the disease will be present in our cohort. If so, the direction of the estimates for deadly diseases strongly related to any blood group will have been lowered or even flipped, relative to any causal relationship. The study design, however, enabled 41-year of follow-up and was deemed reasonable because the blood groups have not been associated with mortality differences. Moreover, the blood group distribution in our cohort was found to be almost identical to a reference population of 2.2 million Danish blood donors. Further, we replicated several findings of associations between the blood groups and severe diseases, including pancreatic cancer.^2, 3^ This may indicate that the potential bias was less prevalent. Further, by controlling for year of birth, the potential effects of immortal time bias were likely reduced, however, this could not be tested. Immortal time biases are potentially applicable in many biobanks studies, e.g. when using the UK Biobank for retrospective studies.^29^

The generalizability of our findings is limited further because our cohort solely included hospitalized patients with known ABO and RhD blood groups. These are patients whom the treating doctor has deemed likely to potentially require a blood transfusion during hospitalization. The patients under study might therefore suffer from other diseases than patients without a determined blood group, and than never hospitalized individuals. Further, diseases that do not require hospitalization could not be examined. Lastly, it was not possible to adjust for possible confounding from the geographical distribution or ethnicity of the patients.^1^

In conclusion, we found the ABO/RhD blood groups to be associated with a wide spectrum of diseases, including cardiovascular-, infectious-, gastrointestinal- and musculoskeletal diseases. This may indicate that some of the selective pressure on the blood groups can be attributed to disease susceptibility differences. We found few associations between the blood groups and age of disease onset.

## Data Availability

This is a register-based study and informed consent for such studies is waived by the Danish Data Protection Agency. Data access was approved by the Danish Patient Safety Authority (3-3013-1731), the Danish Data Protection Agency (DT SUND 2016-50 and 2017-57) and the Danish Health Data Authority (FSEID 00003092 and FSEID 00003724). The study was conducted using anonymized patient data not publicly accessible. Data access may be granted by contacting the Danish Patient Safety Authority, the Danish Data Protection Agency, and the Danish Health Data Authority.

## Acknowledgements

This study was performed as a part of the CAG (Clinical Academic Group) Center for Endotheliomics under the Greater Copenhagen Health Science Partners (GCHSP).

## Data sharing statement

This is a register-based study and informed consent for such studies is waived by the Danish Data Protection Agency. Data access was approved by the Danish Patient Safety Authority (3– 3013–1731), the Danish Data Protection Agency (DT SUND 2016–50 and 2017–57) and the Danish Health Data Authority (FSEID 00003092 and FSEID 00003724). The study was conducted using anonymized patient data not publicly accessible. Data access may be granted by contacting the Danish Patient Safety Authority, the Danish Data Protection Agency, and the Danish Health Data Authority.

## Supplementary Materials

**Figure 1.**
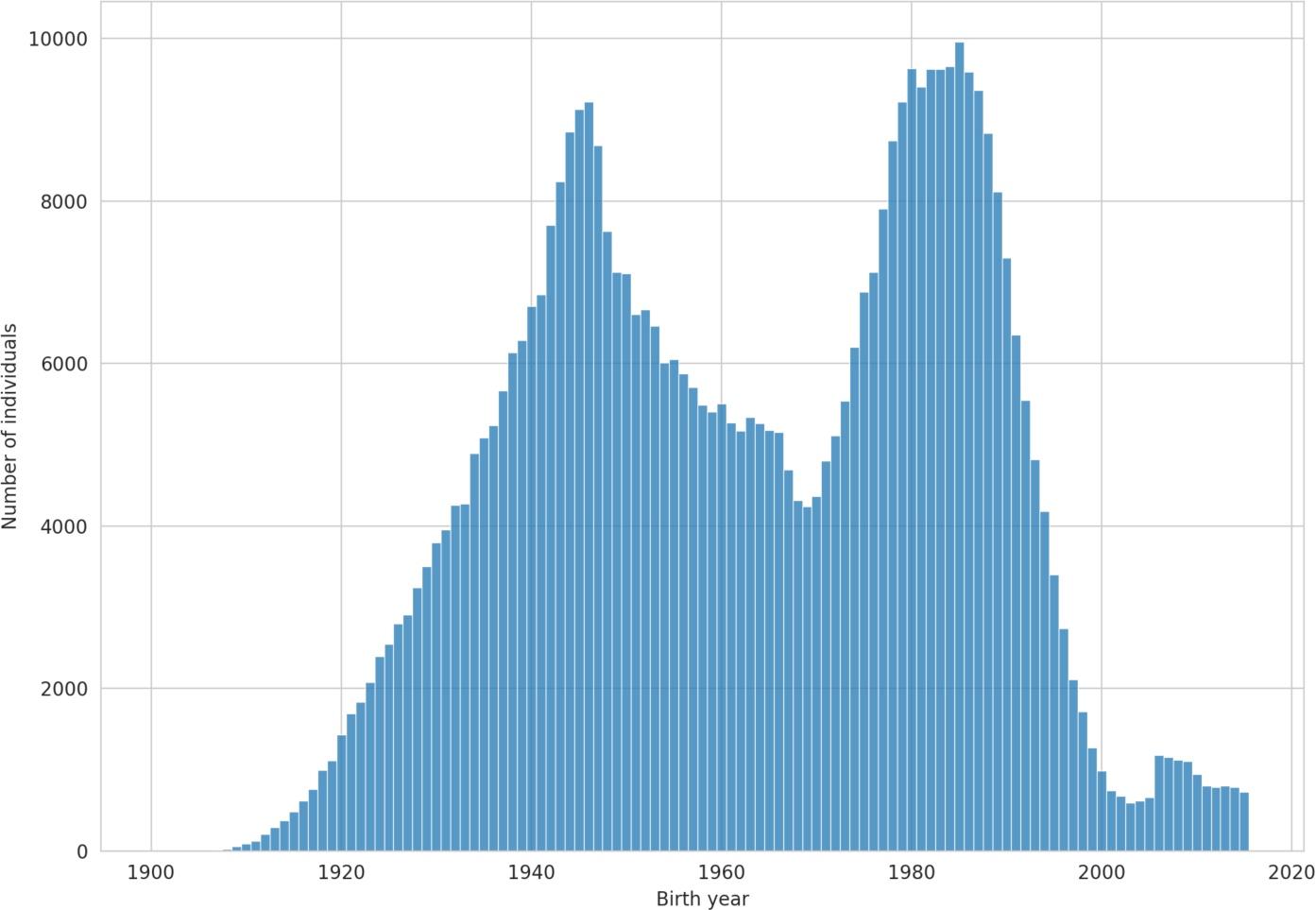
The study sample birth year distribution.

**Table 1.**
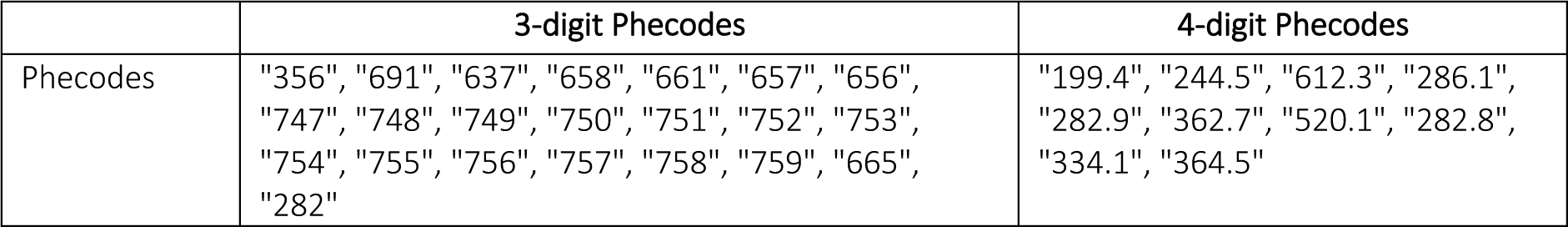
List of Phecodes defined as congenital or hereditary.

|  | 3-digit Phecodes | 4-digit Phecodes |
| --- | --- | --- |
| Phecodes | "356", "691", "637", "658", "661", "657", "656",<br>"747", "748", "749", "750", "751", "752", "753",<br>"754", "755", "756", "757", "758", "759", "665",<br>"282" | "199.4", "244.5", "612.3", "286.1",<br>"282.9", "362.7", "520.1", "282.8",<br>"334.1", "364.5" |

**Table 2:**
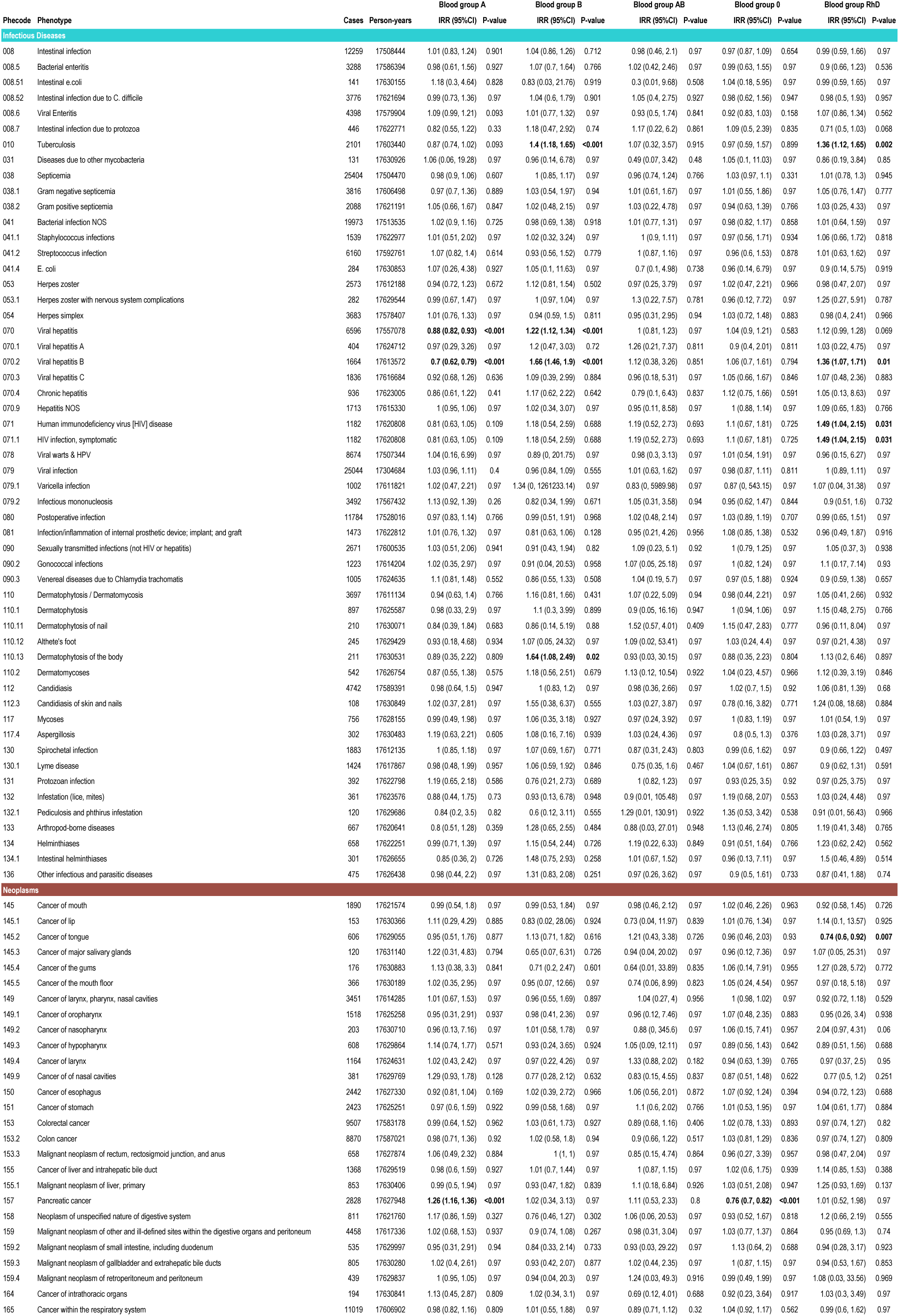
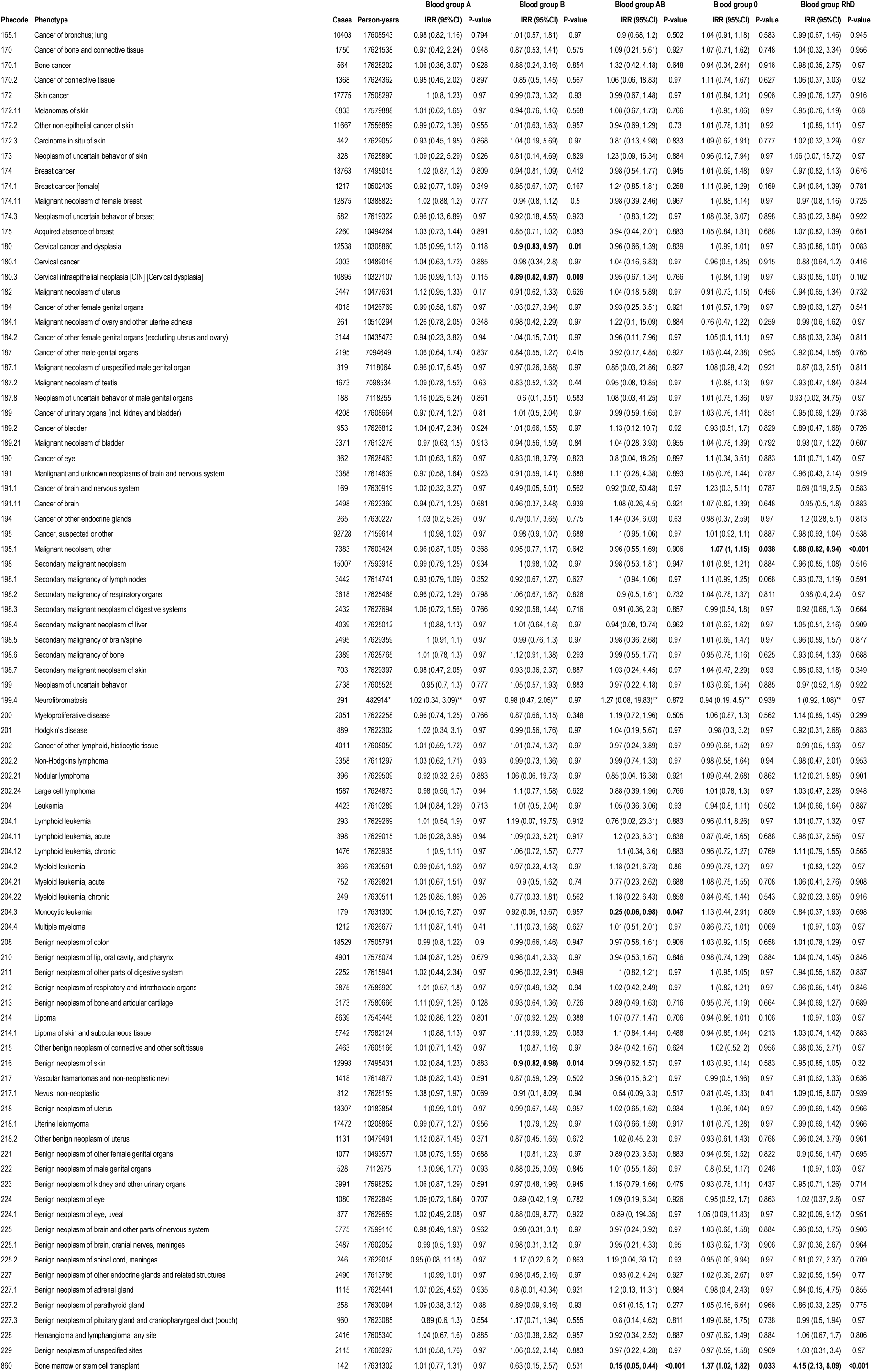
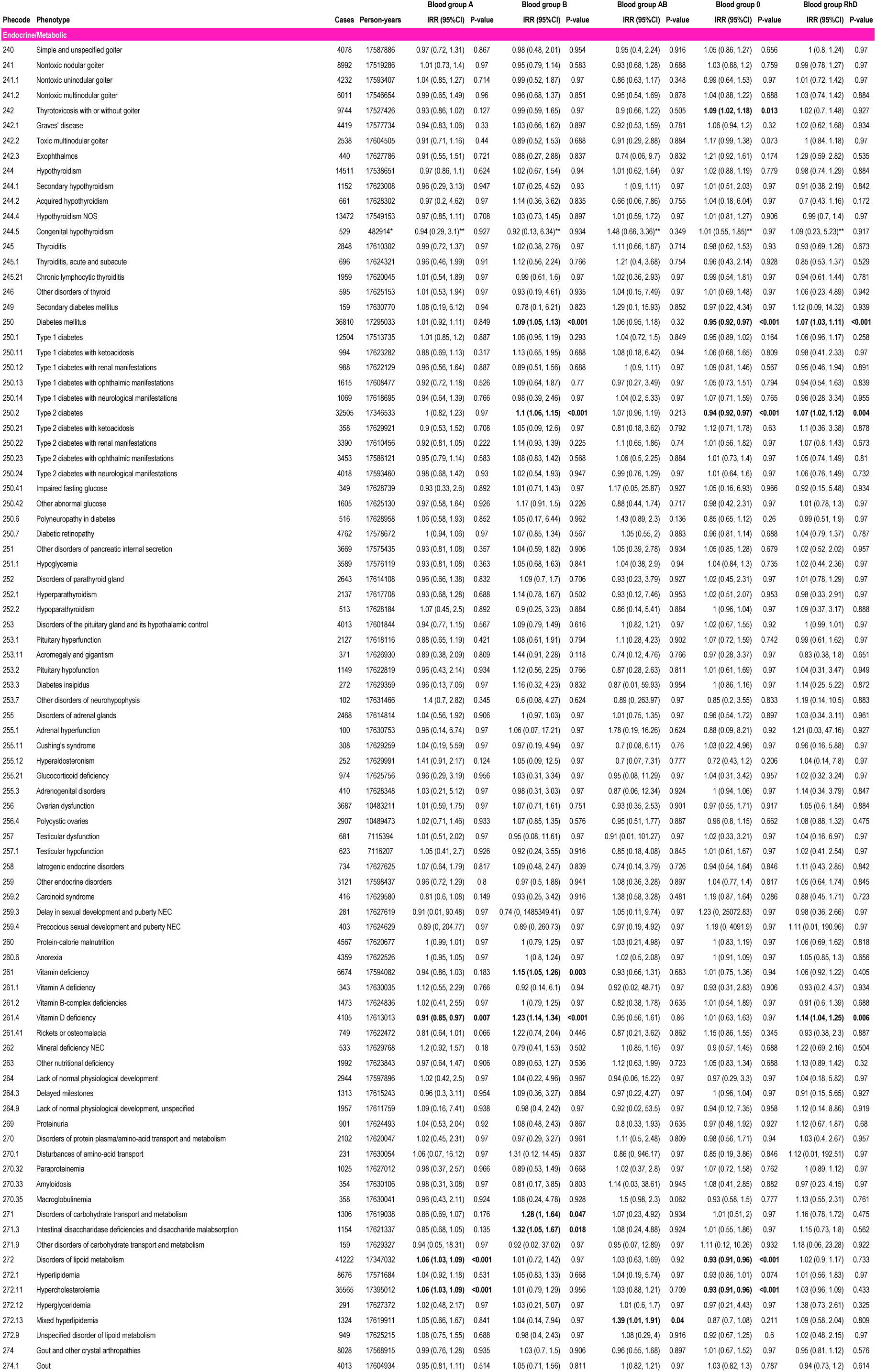
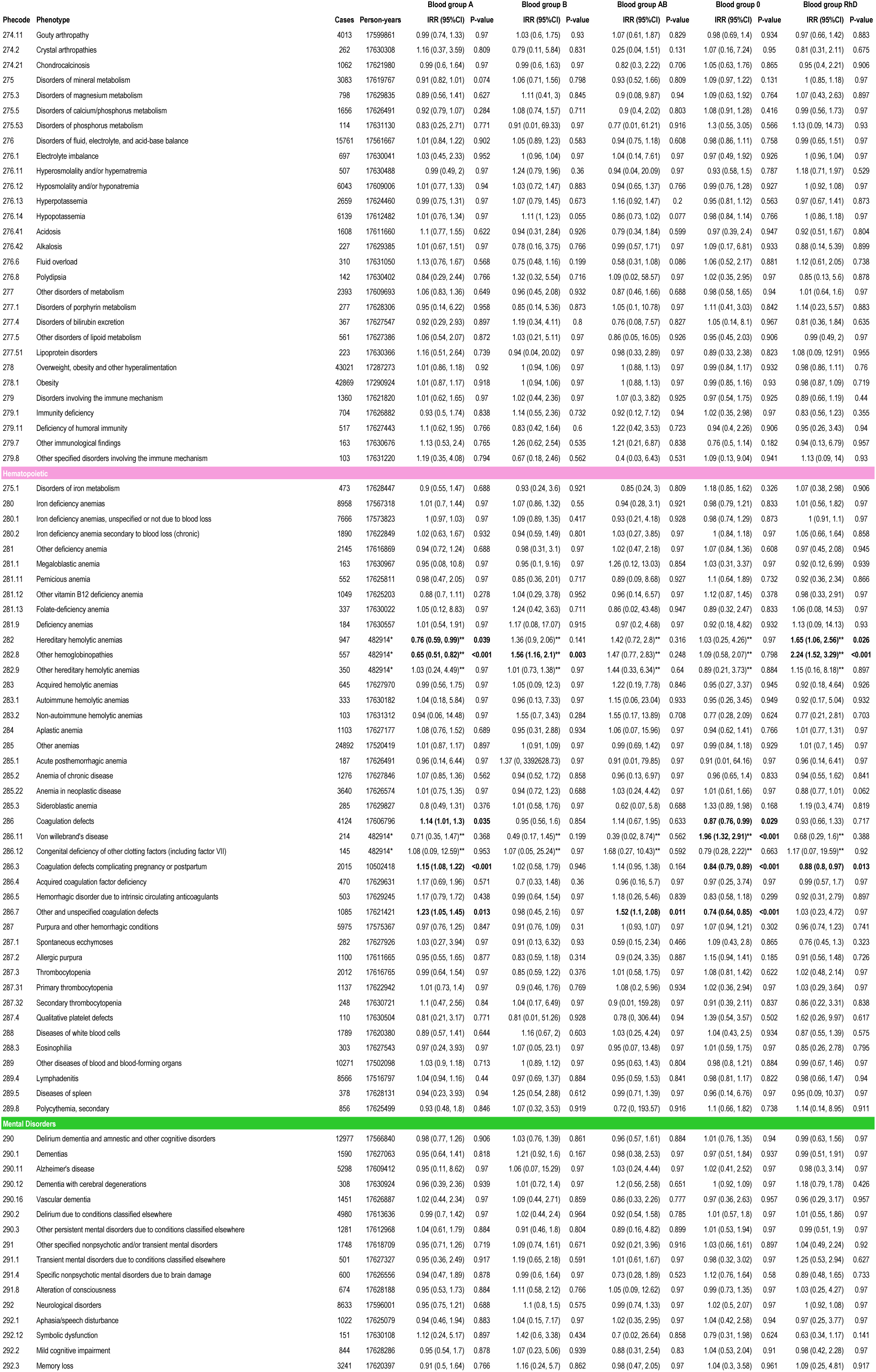
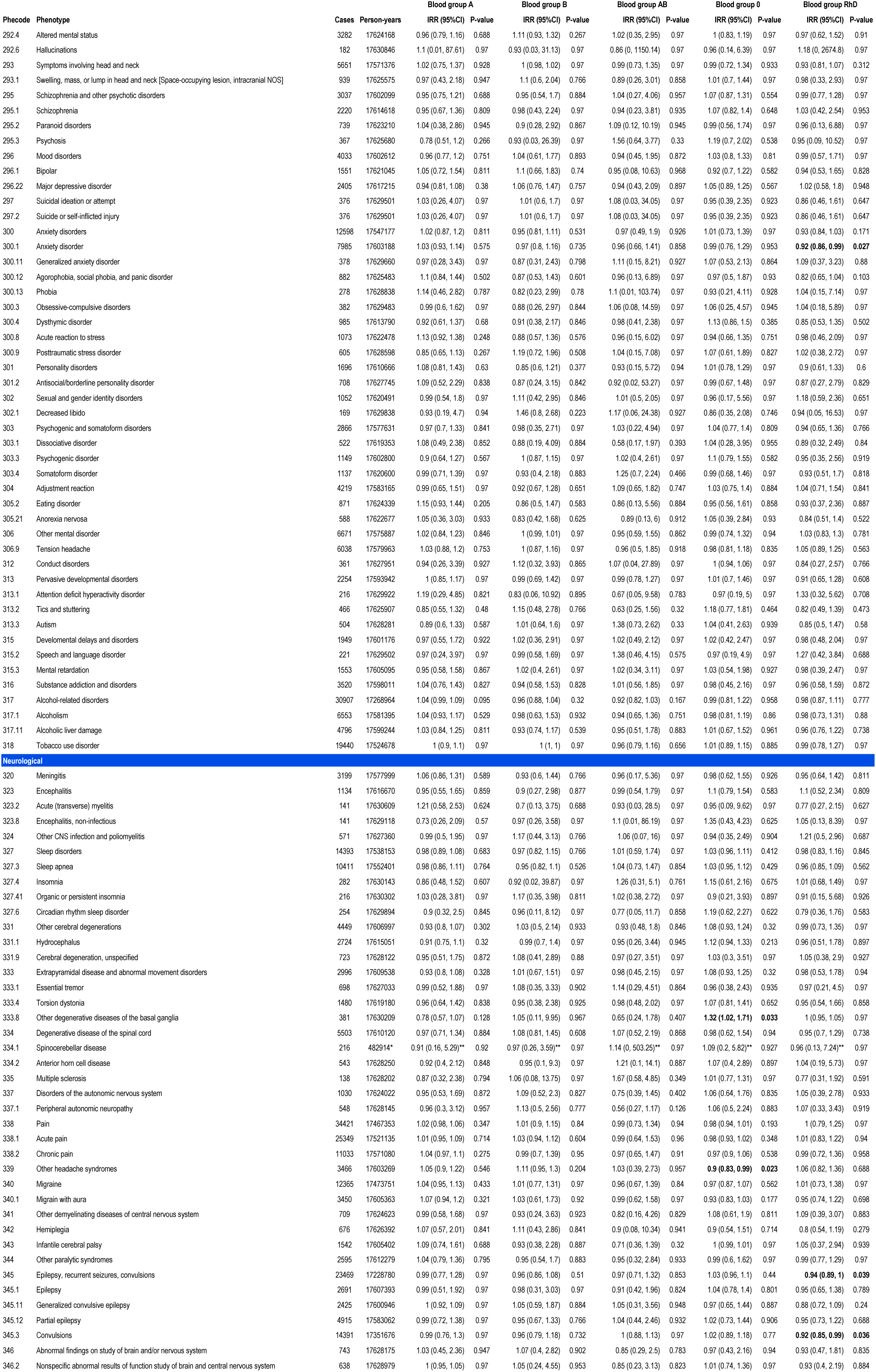
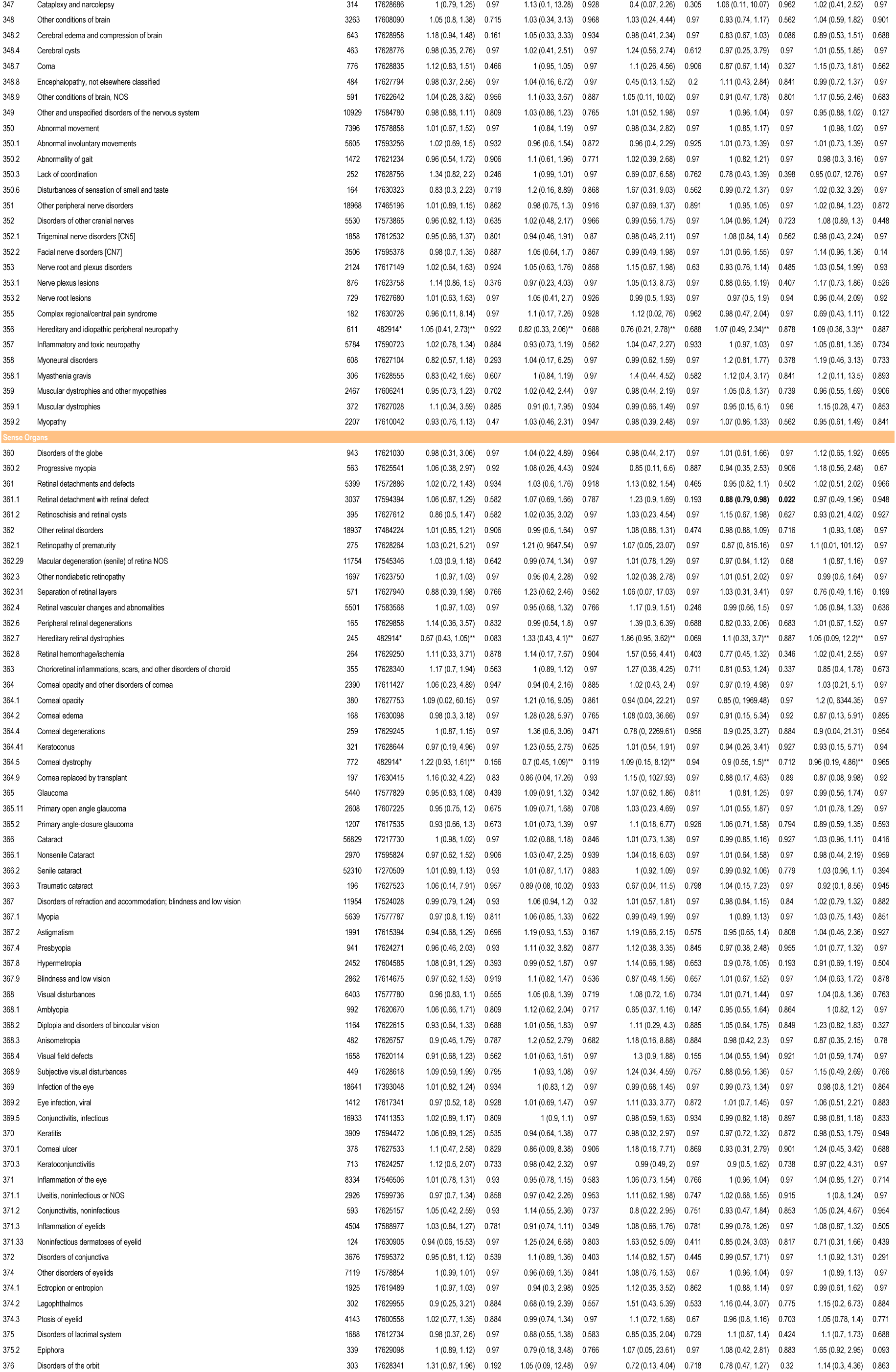
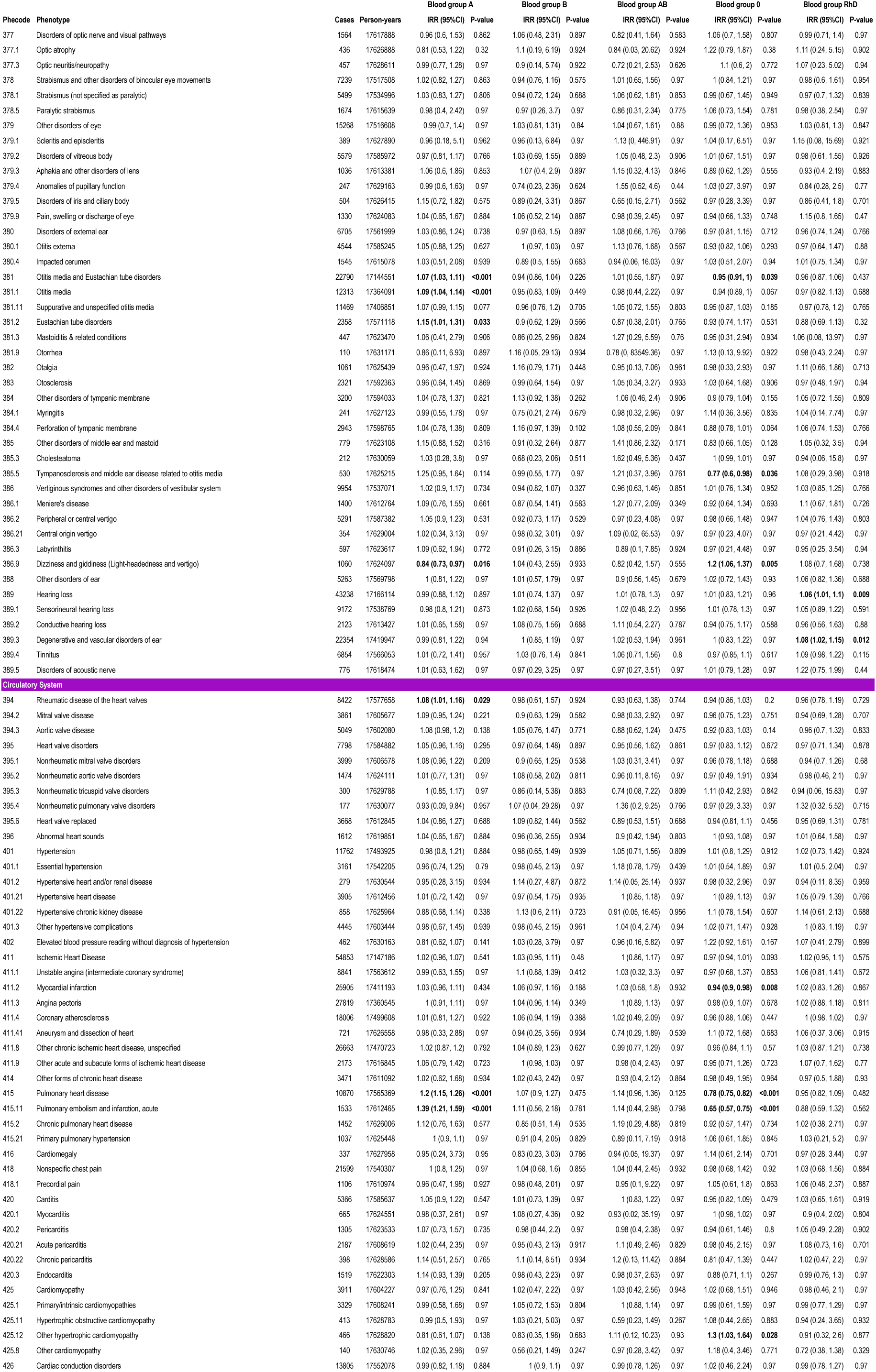
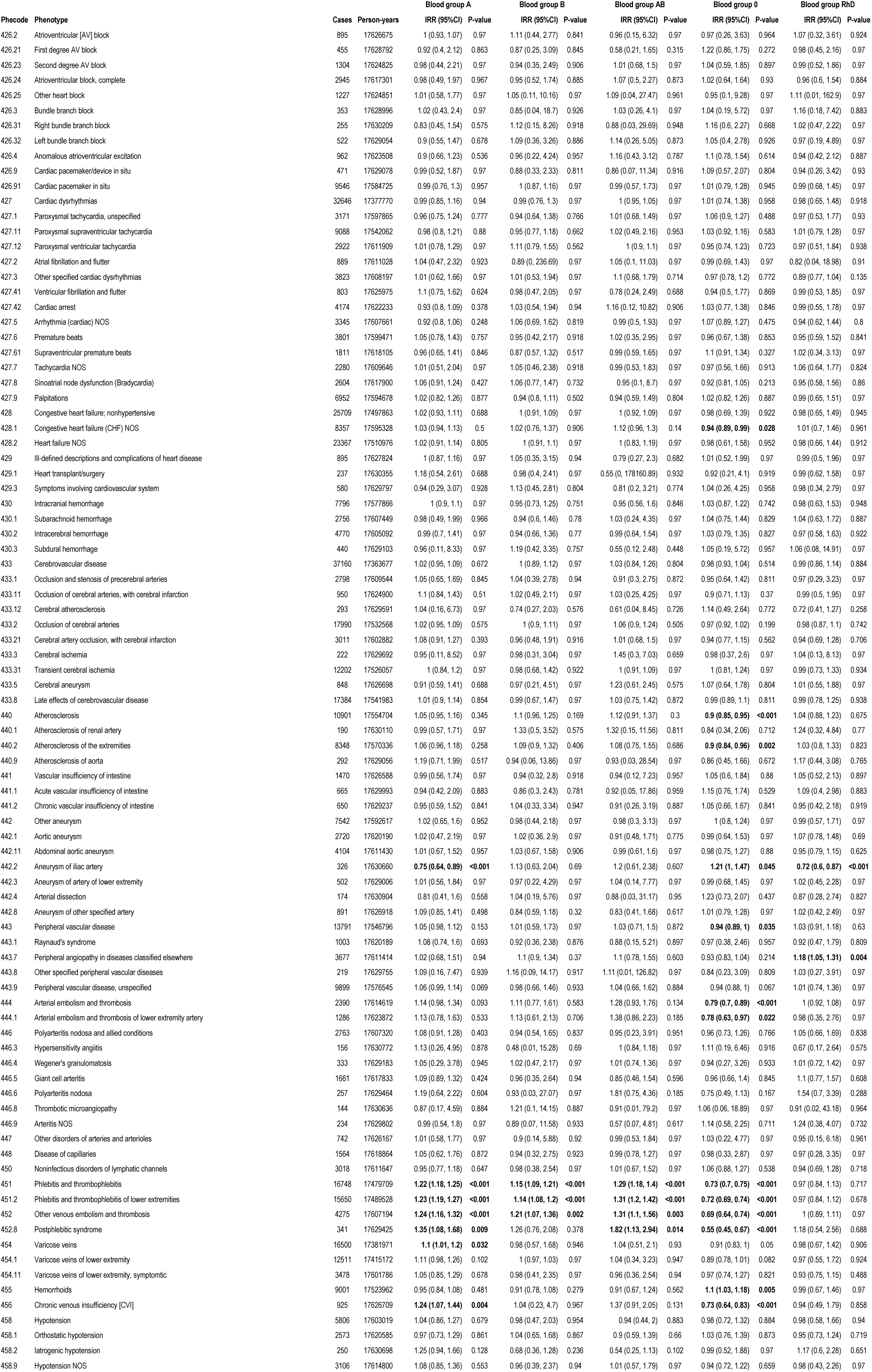
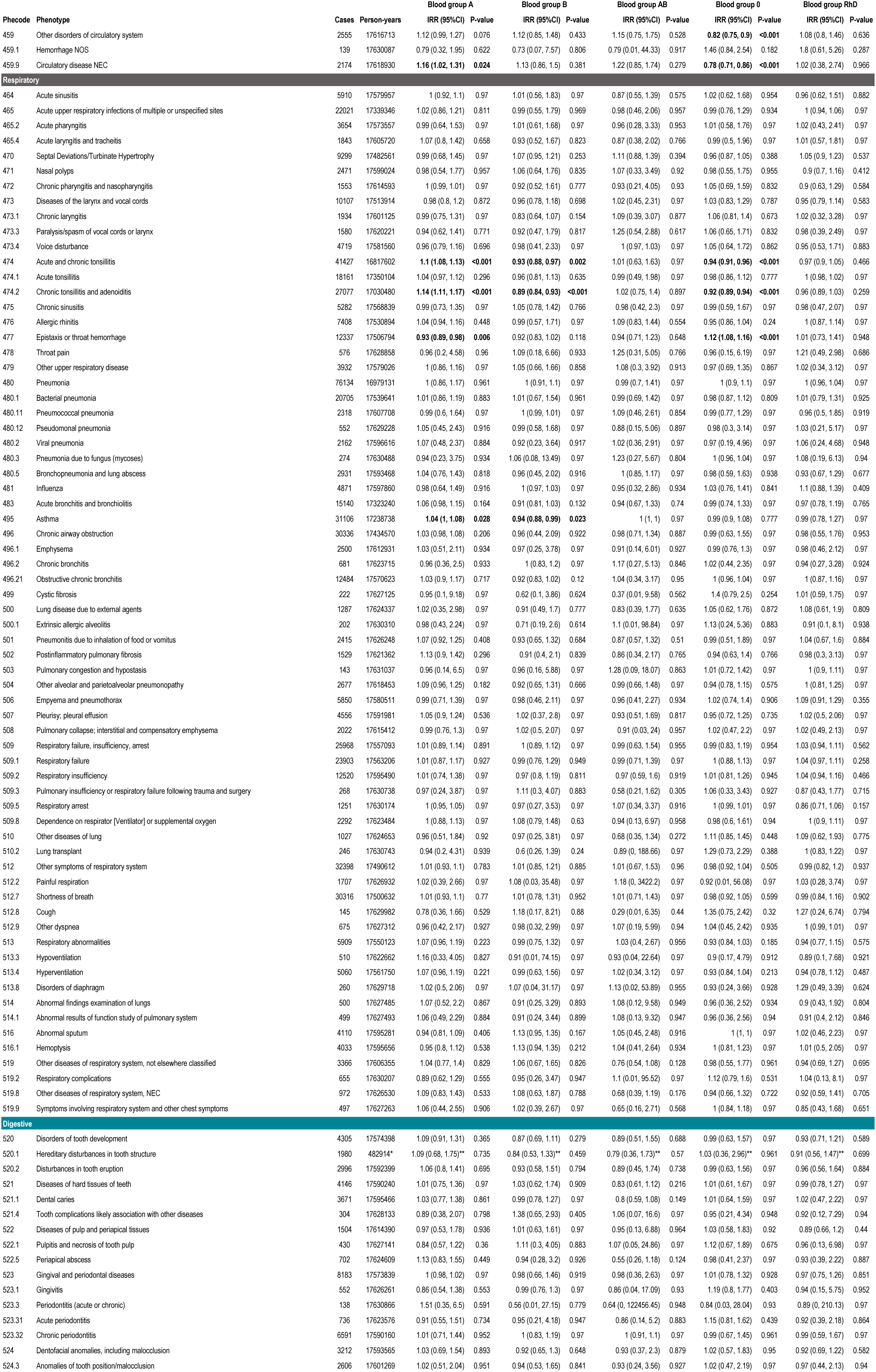
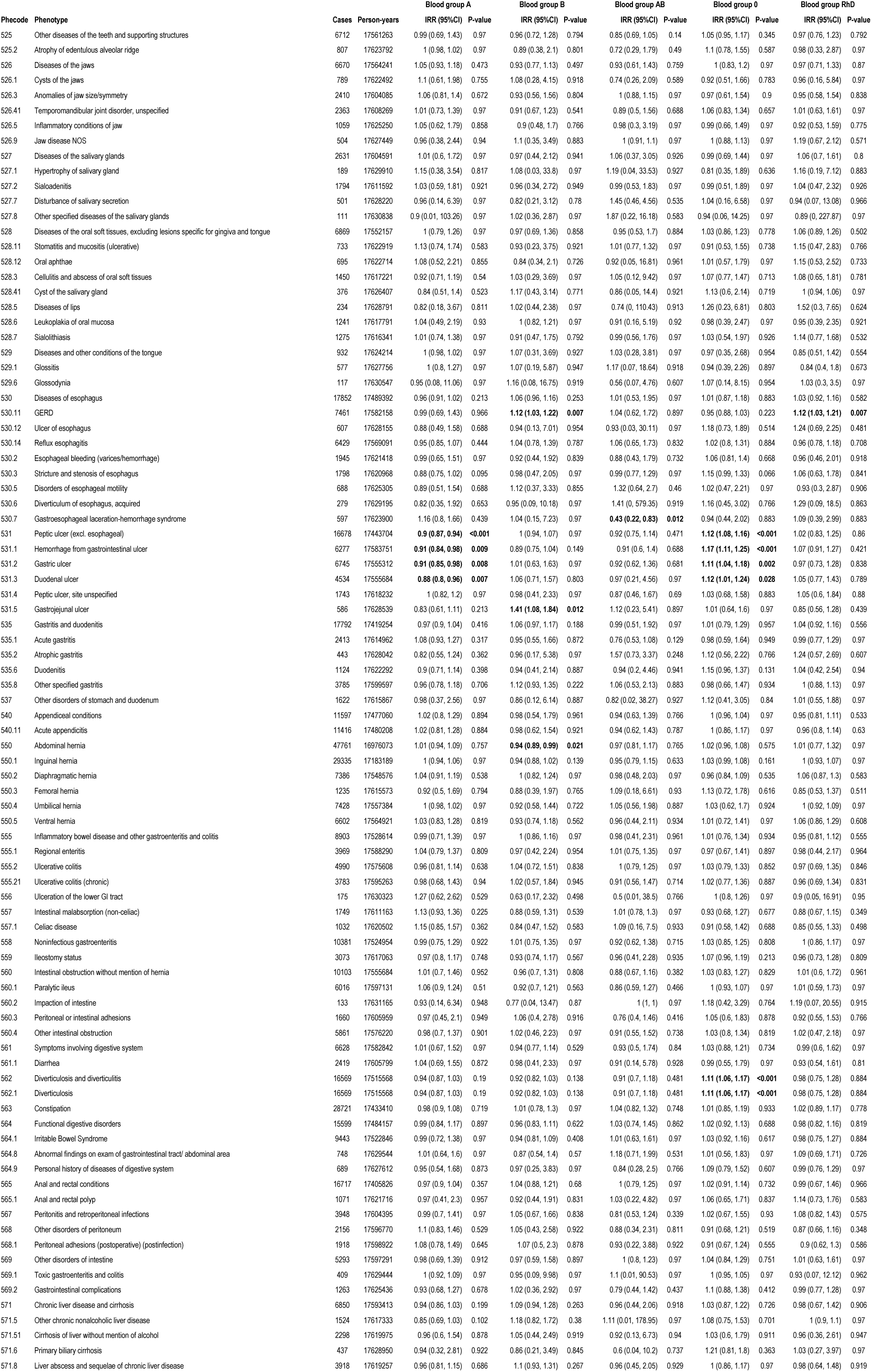
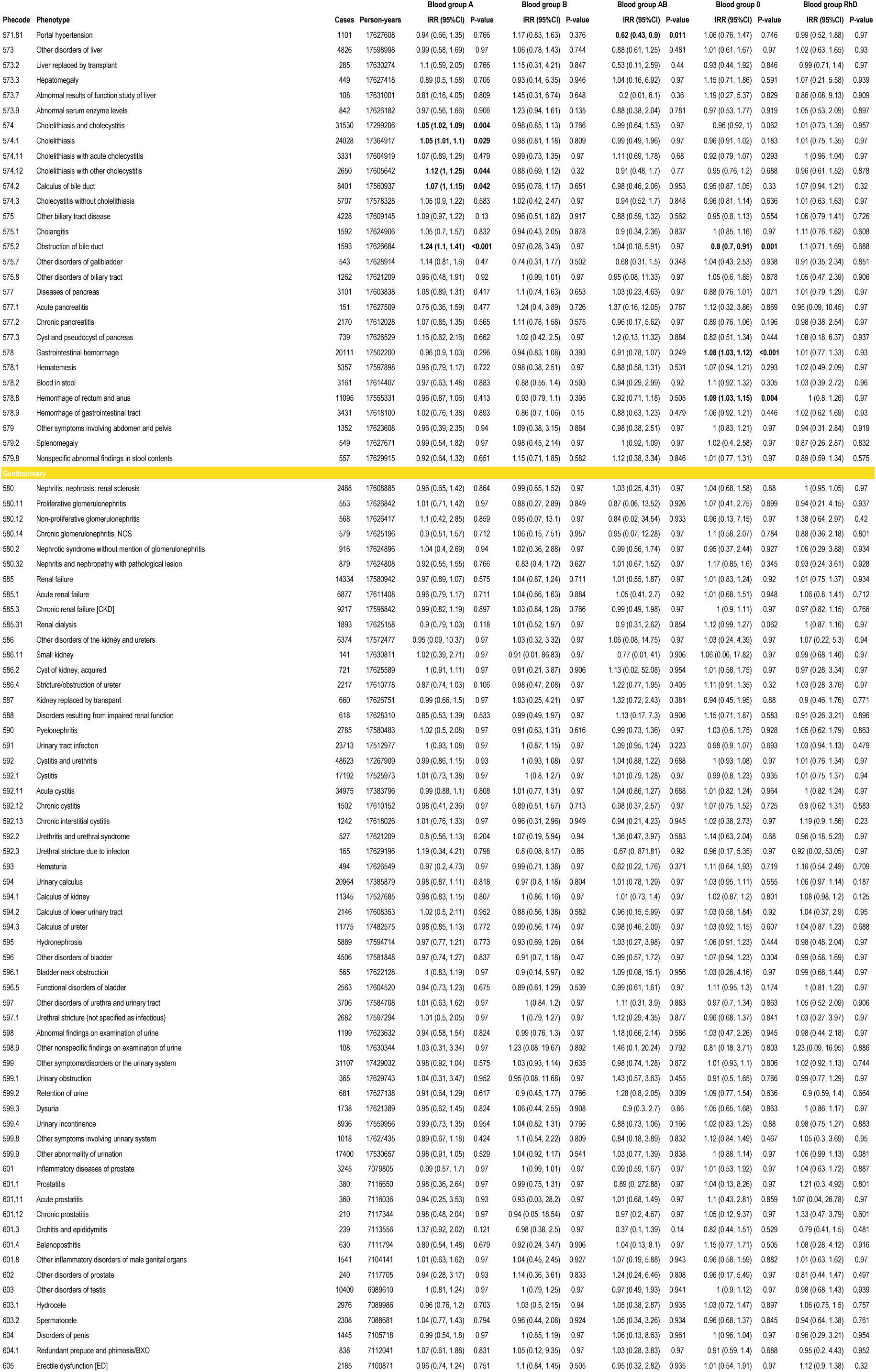
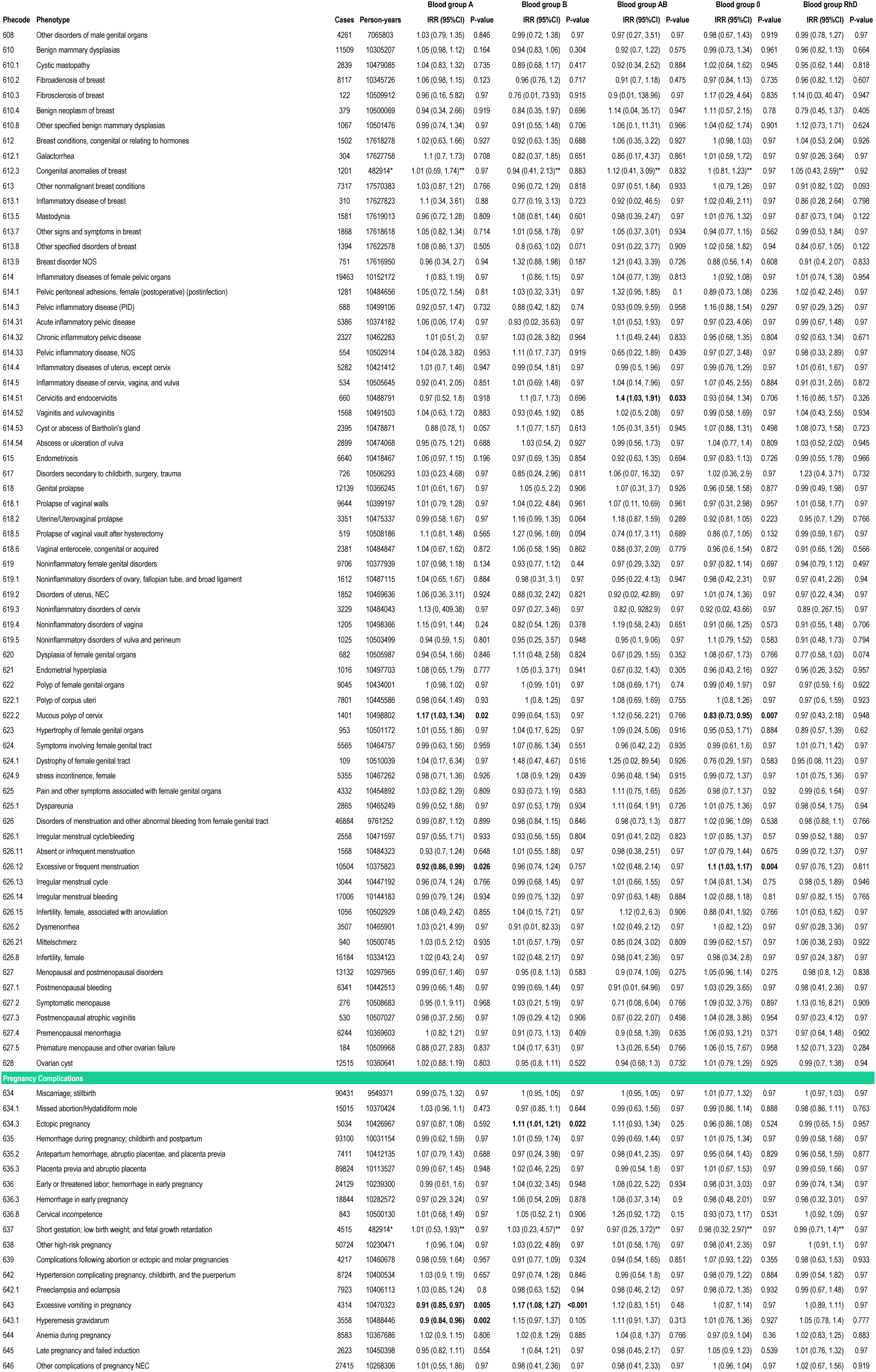
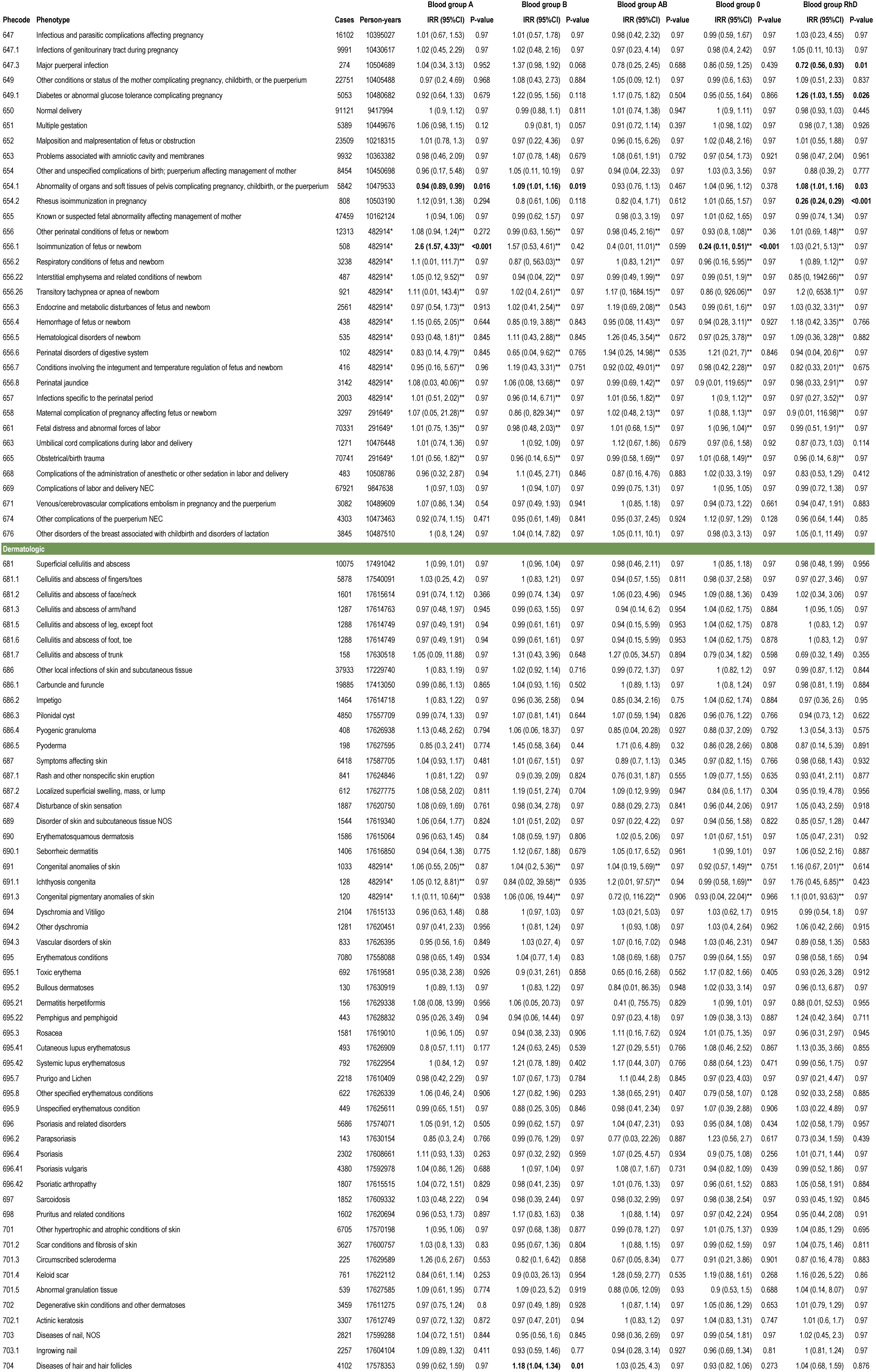
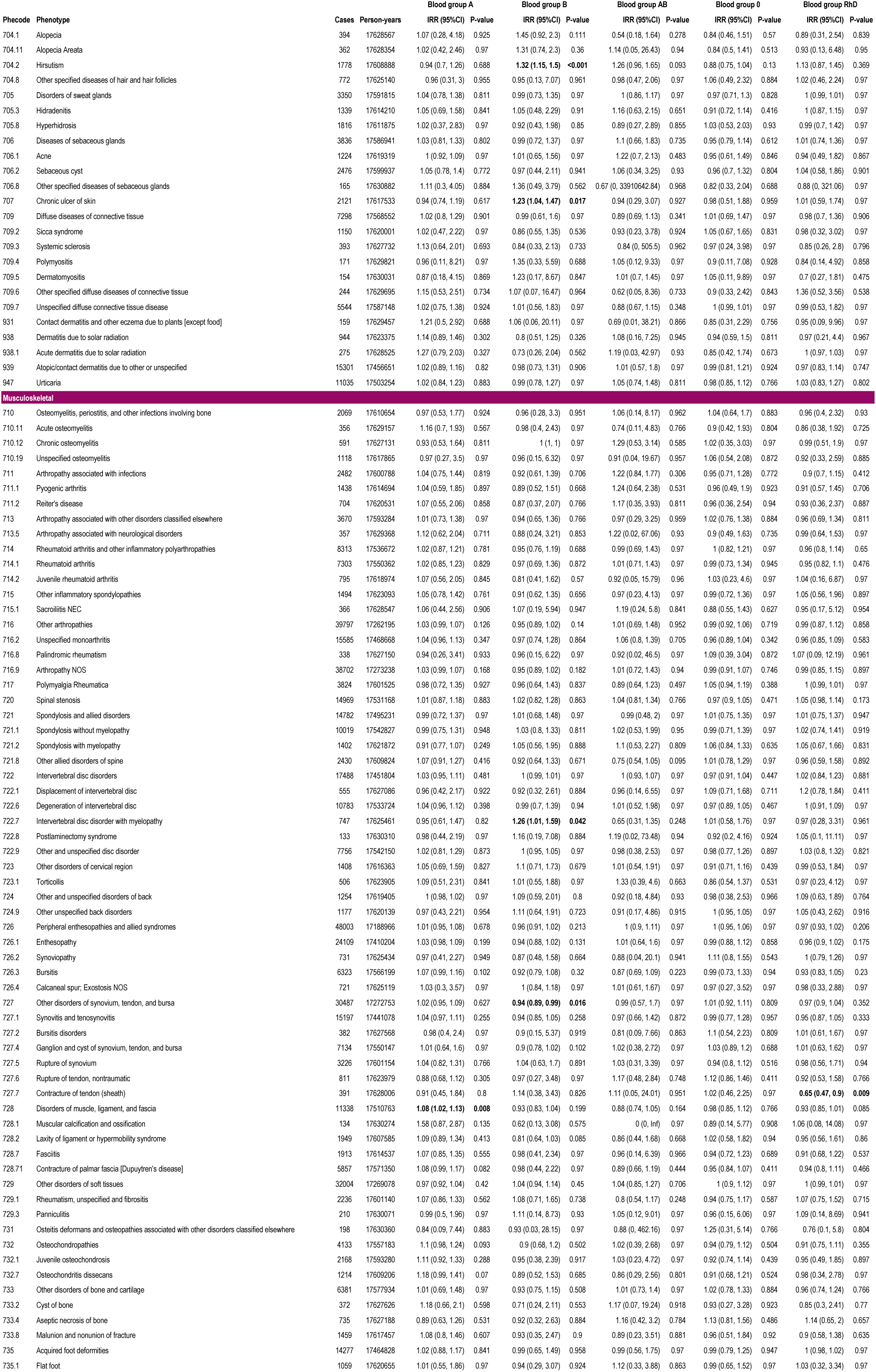
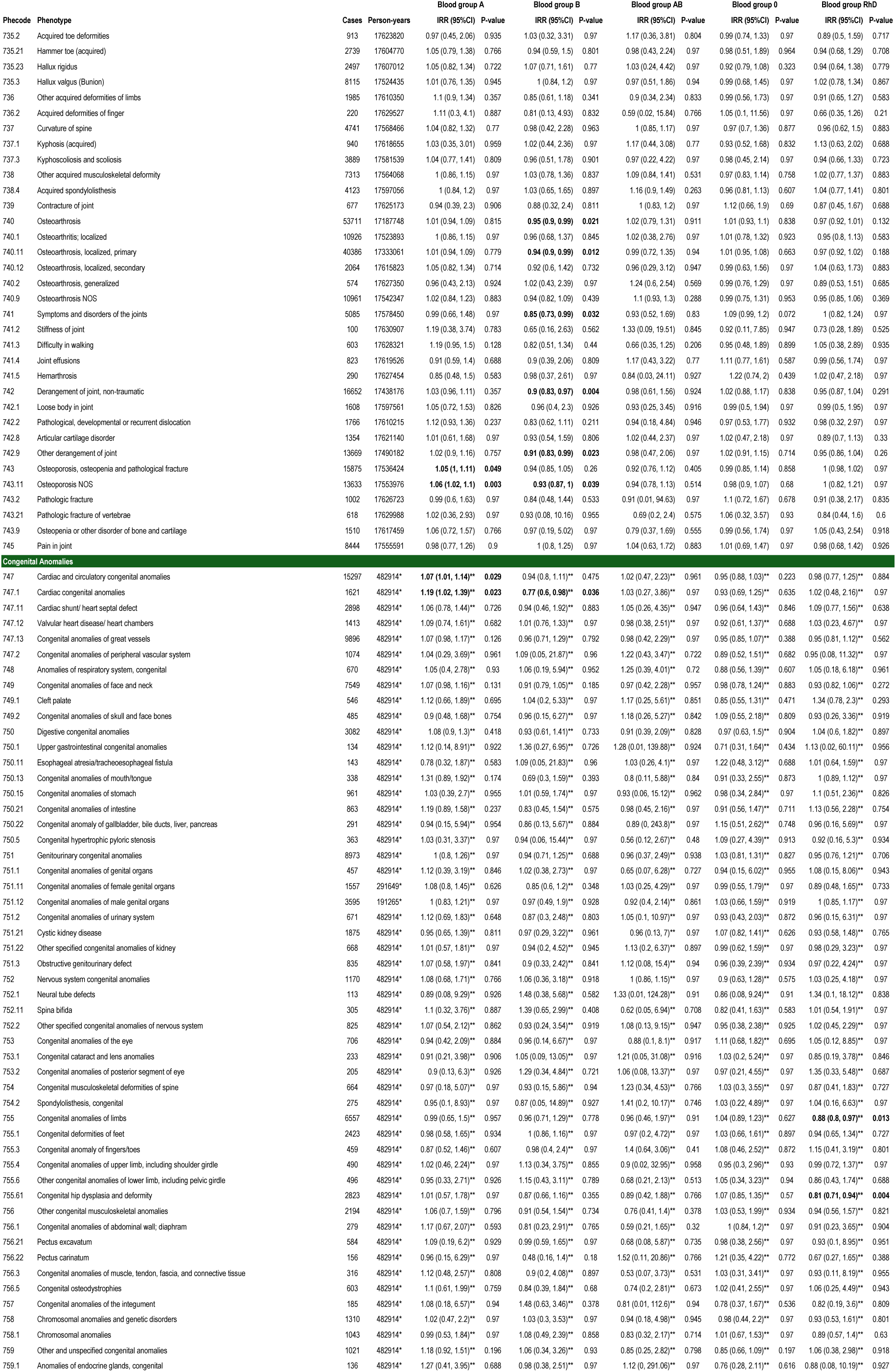
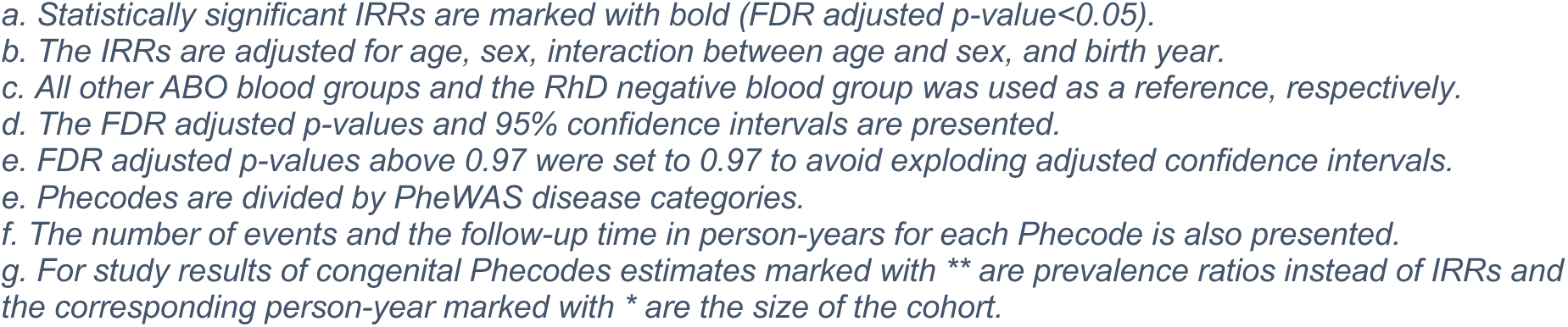
Associations between the ABO/RhD blood groups and Phecode incidence rate ratios of all analyzed Phecodes

**Table 3:**
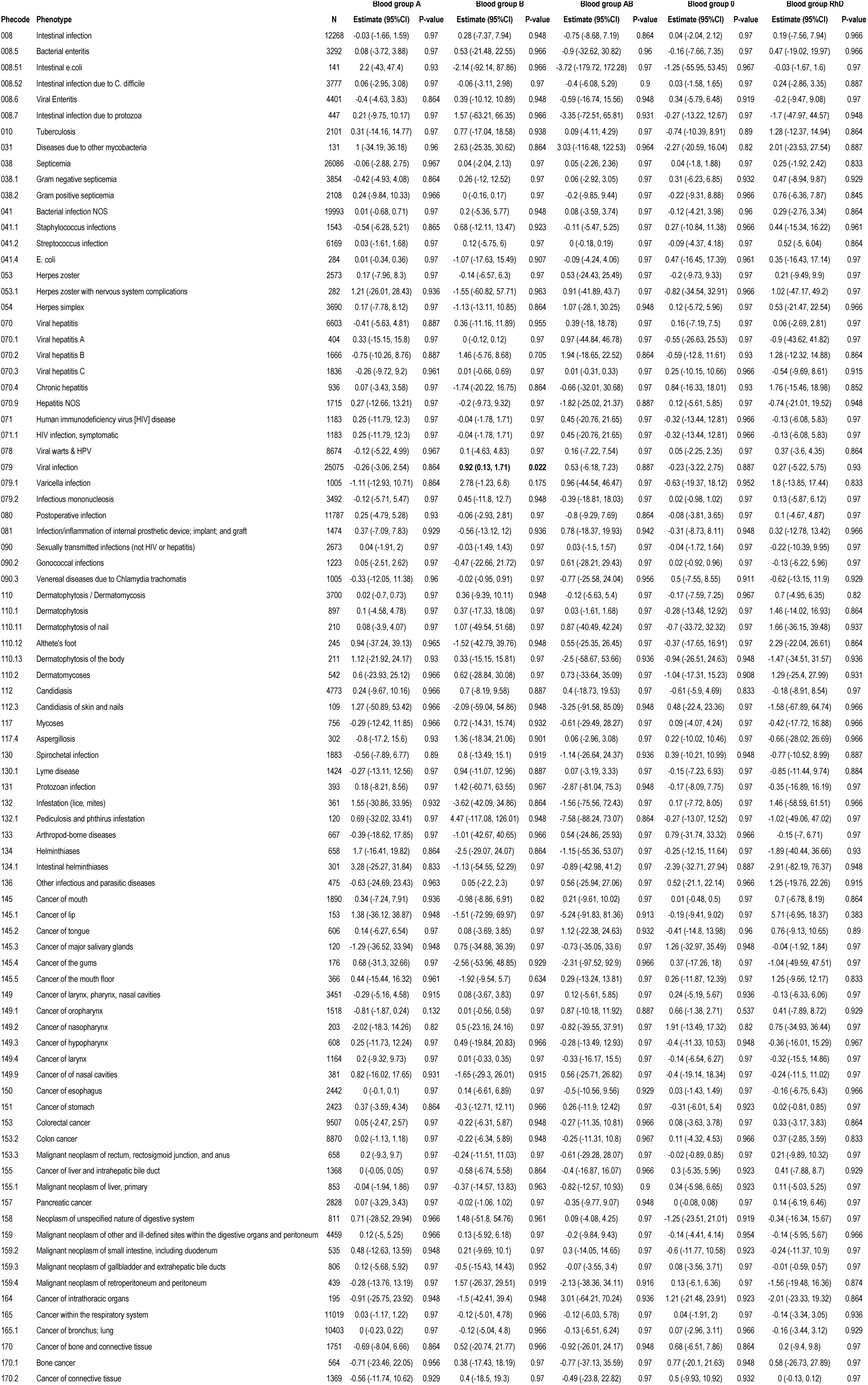
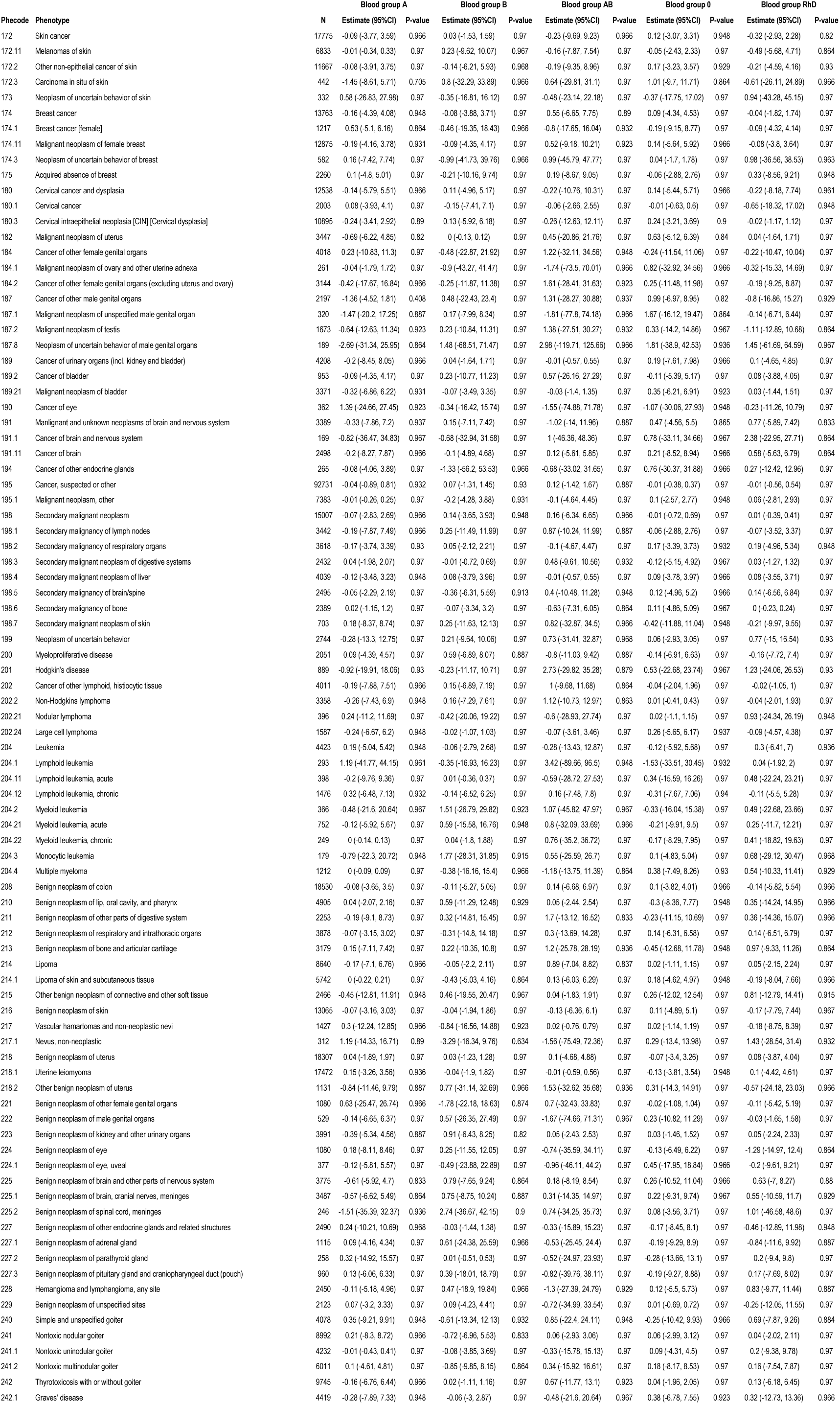
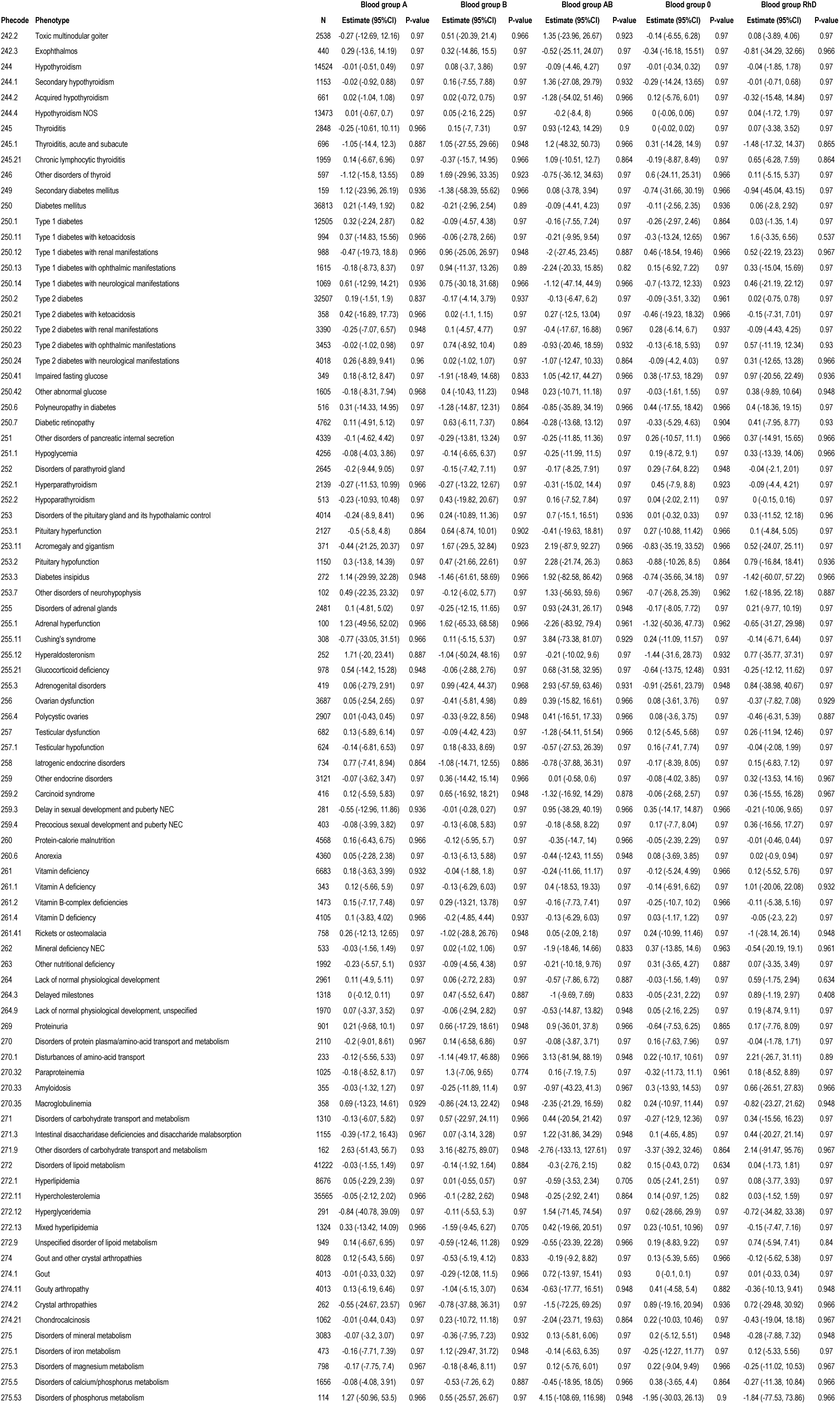
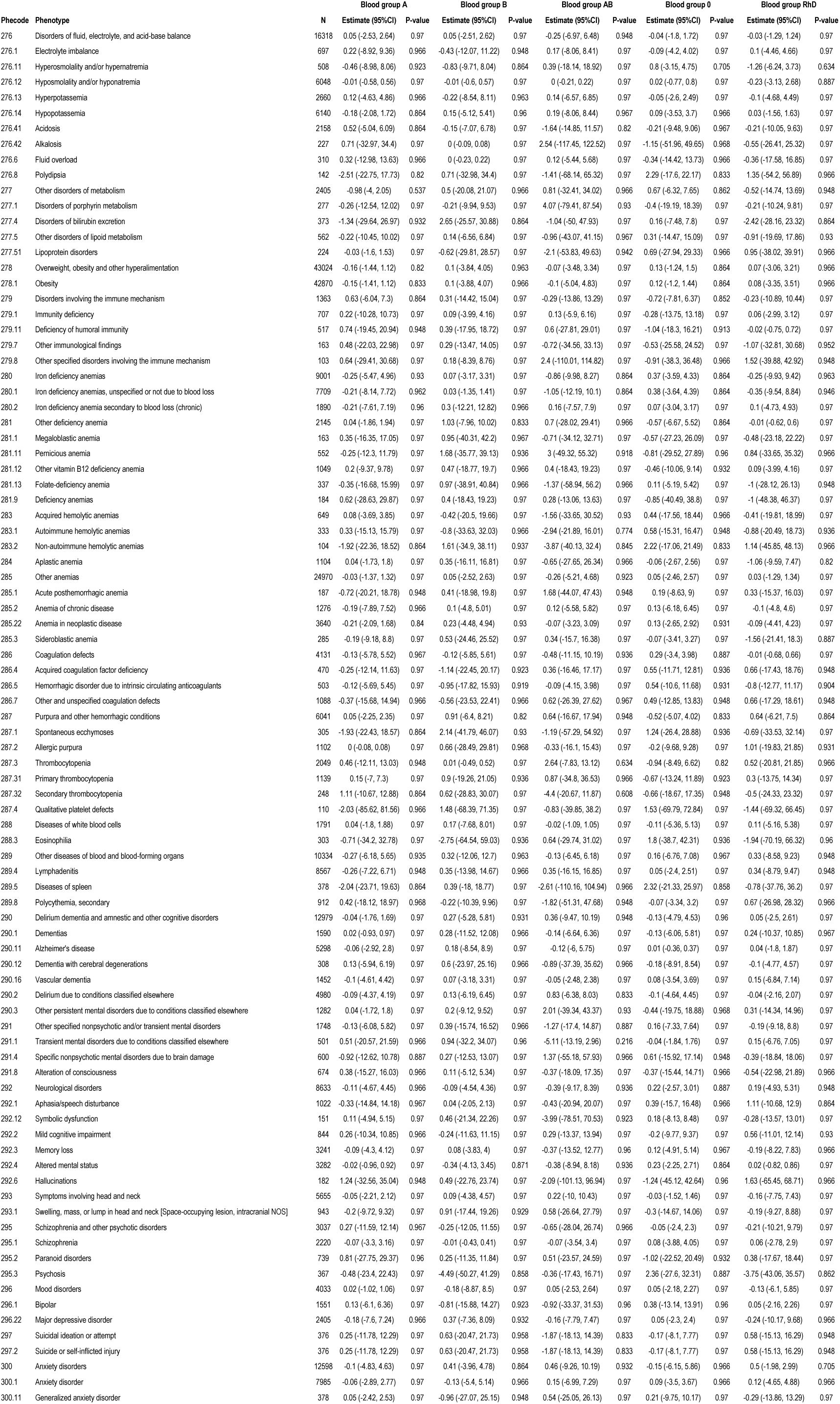
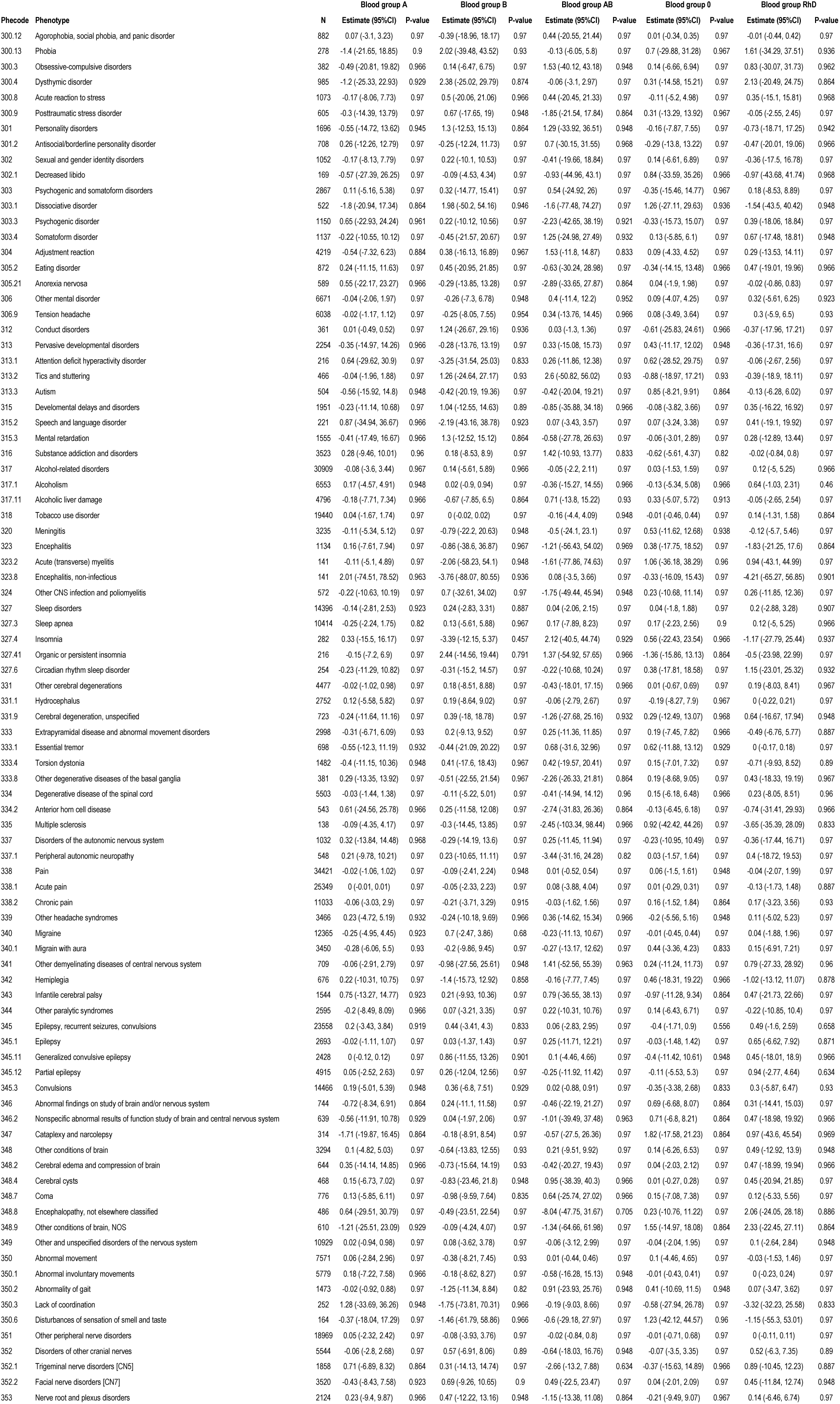
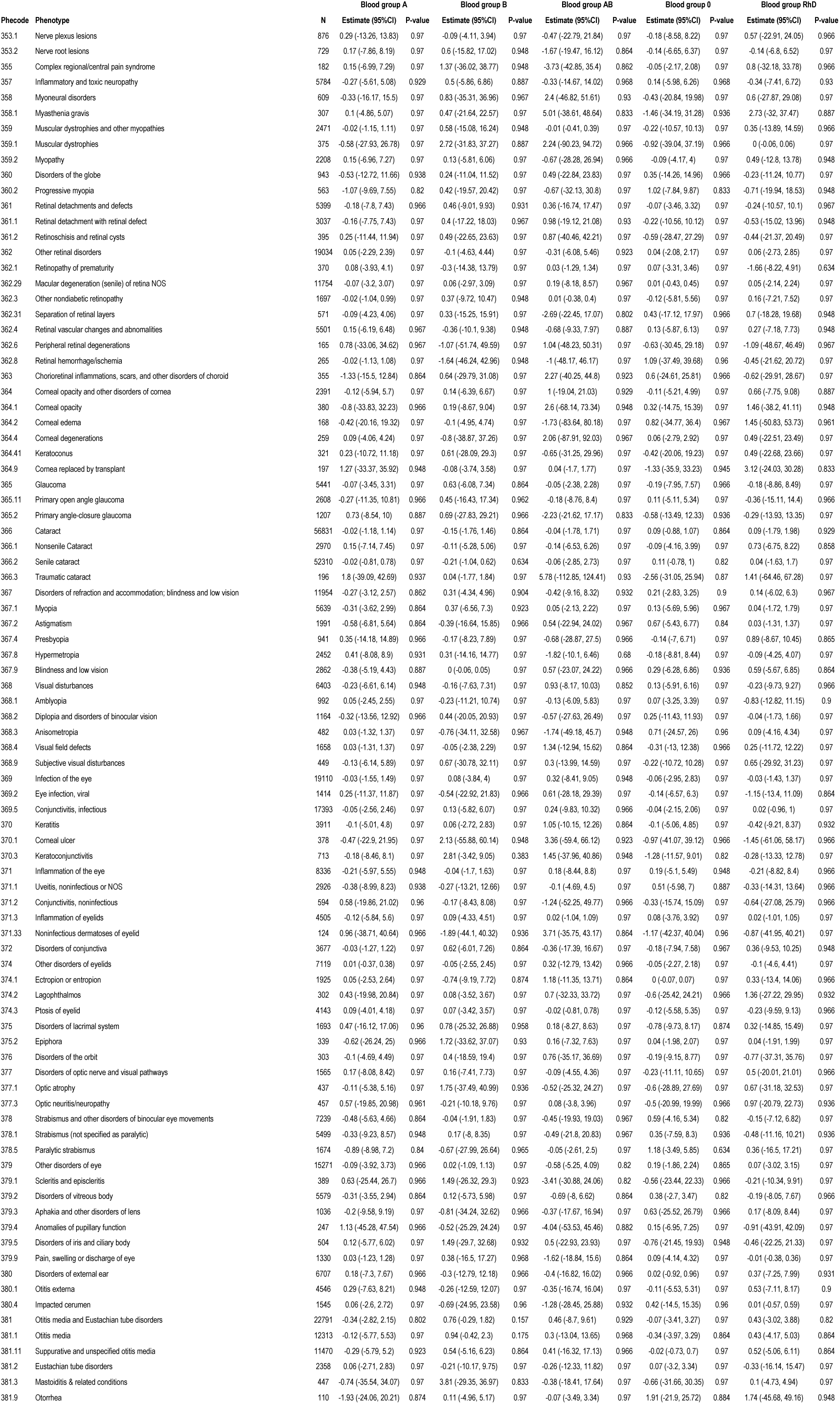
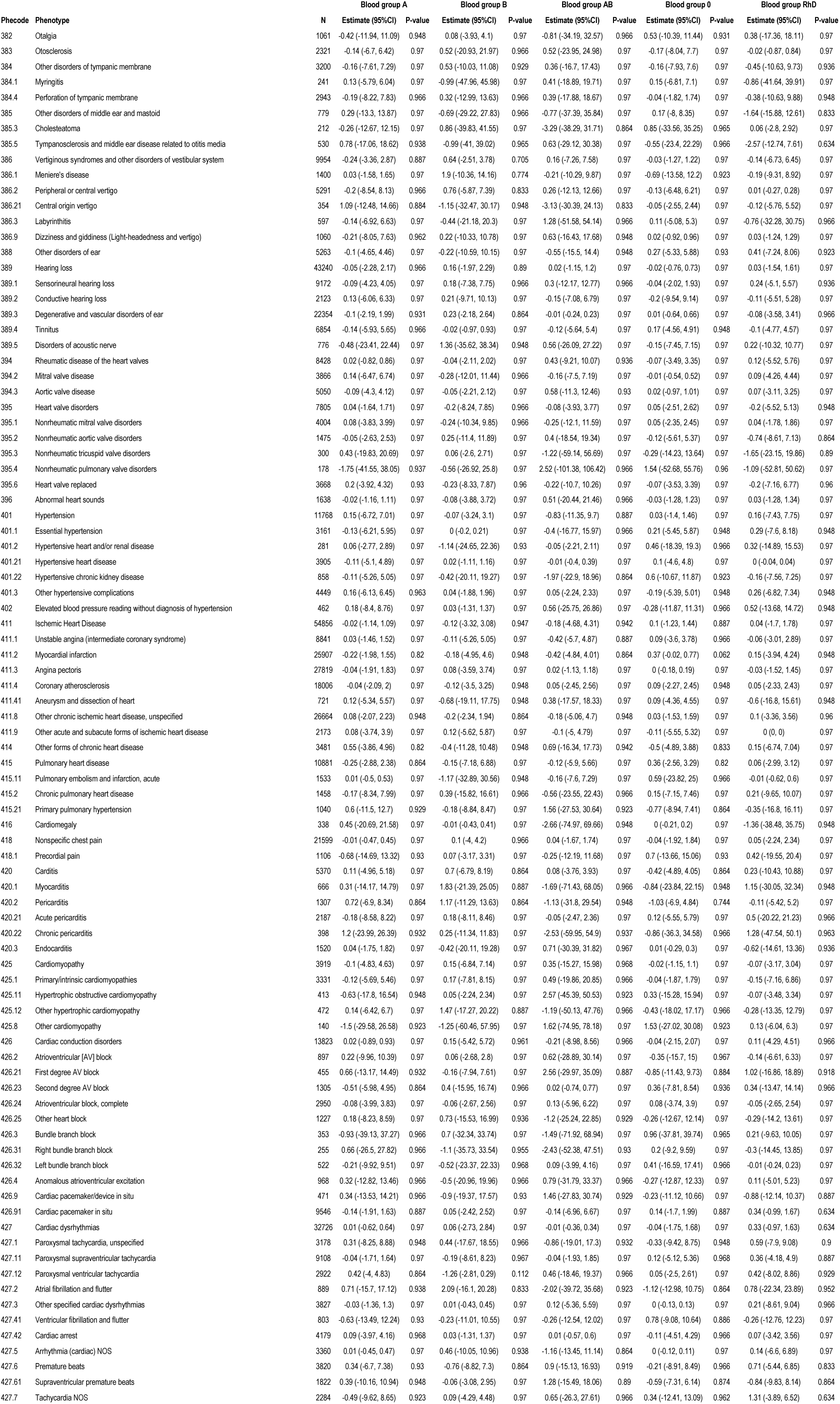
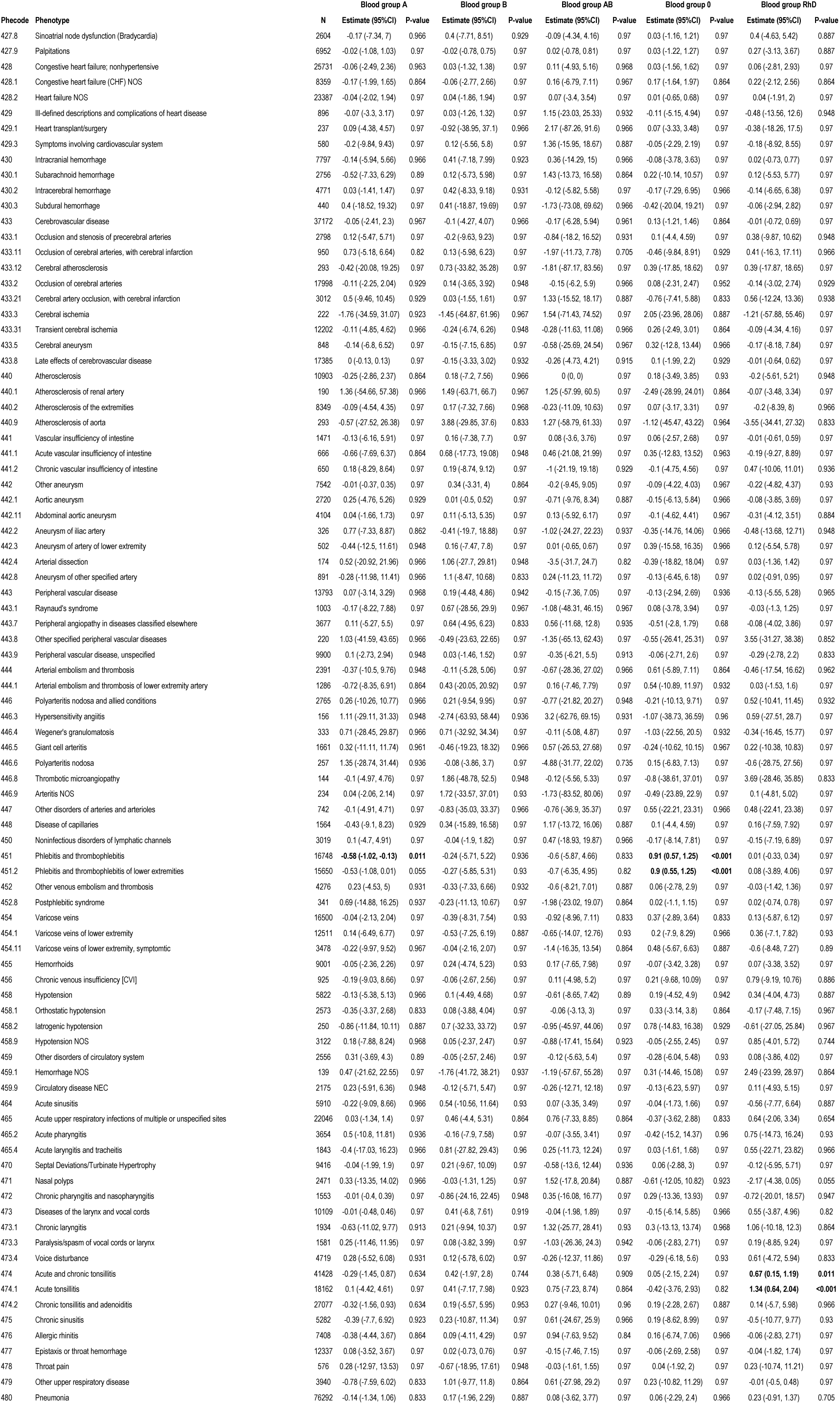
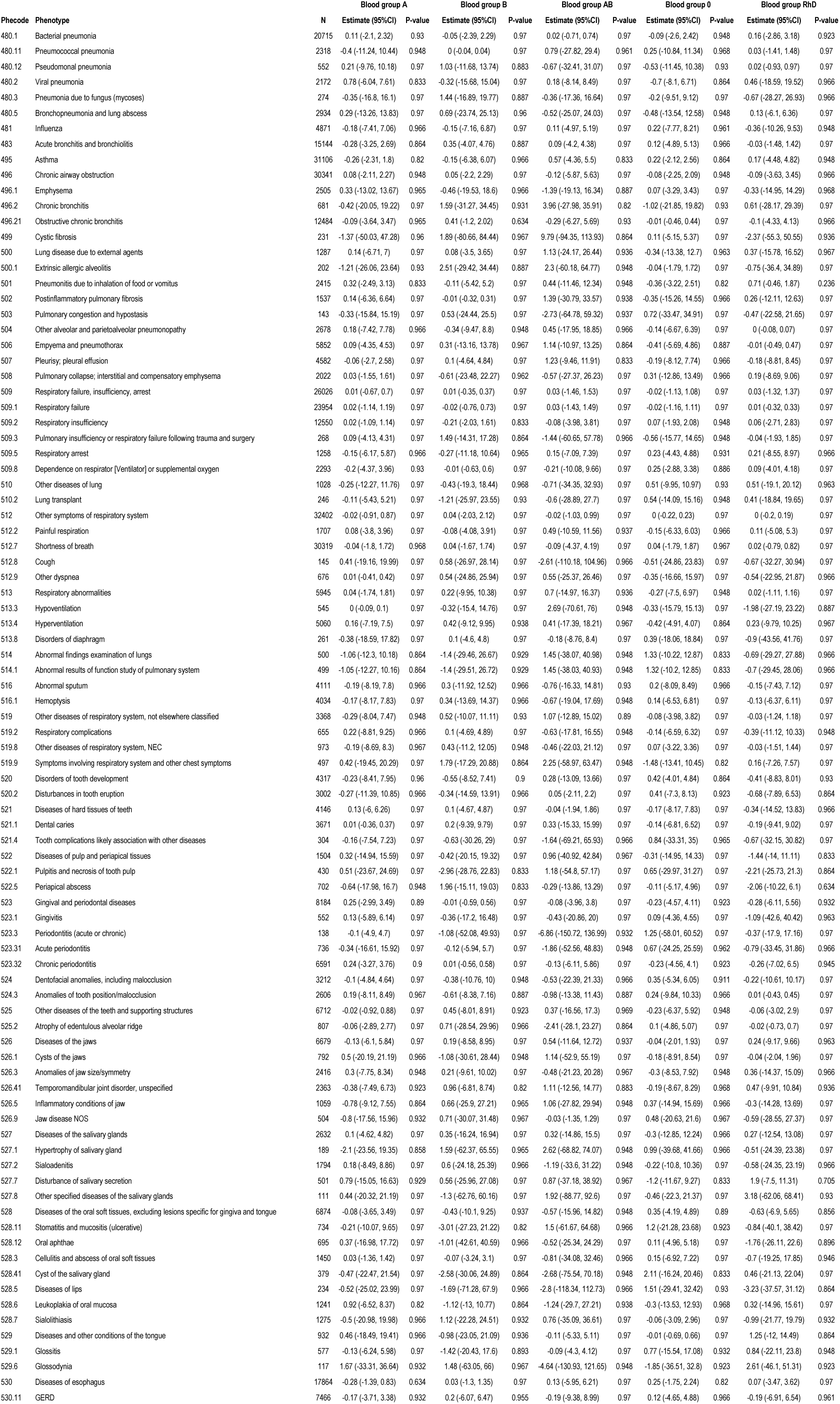
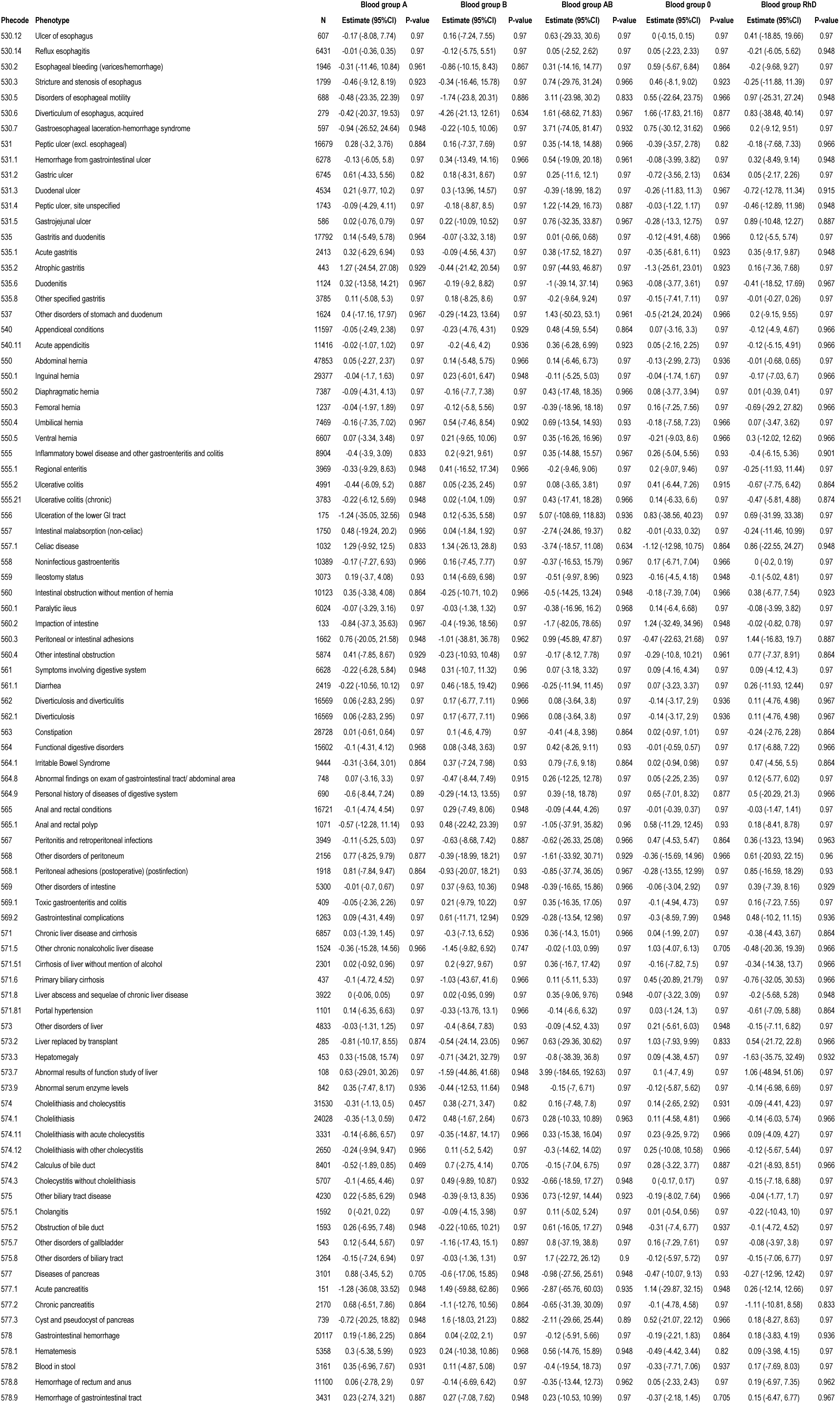
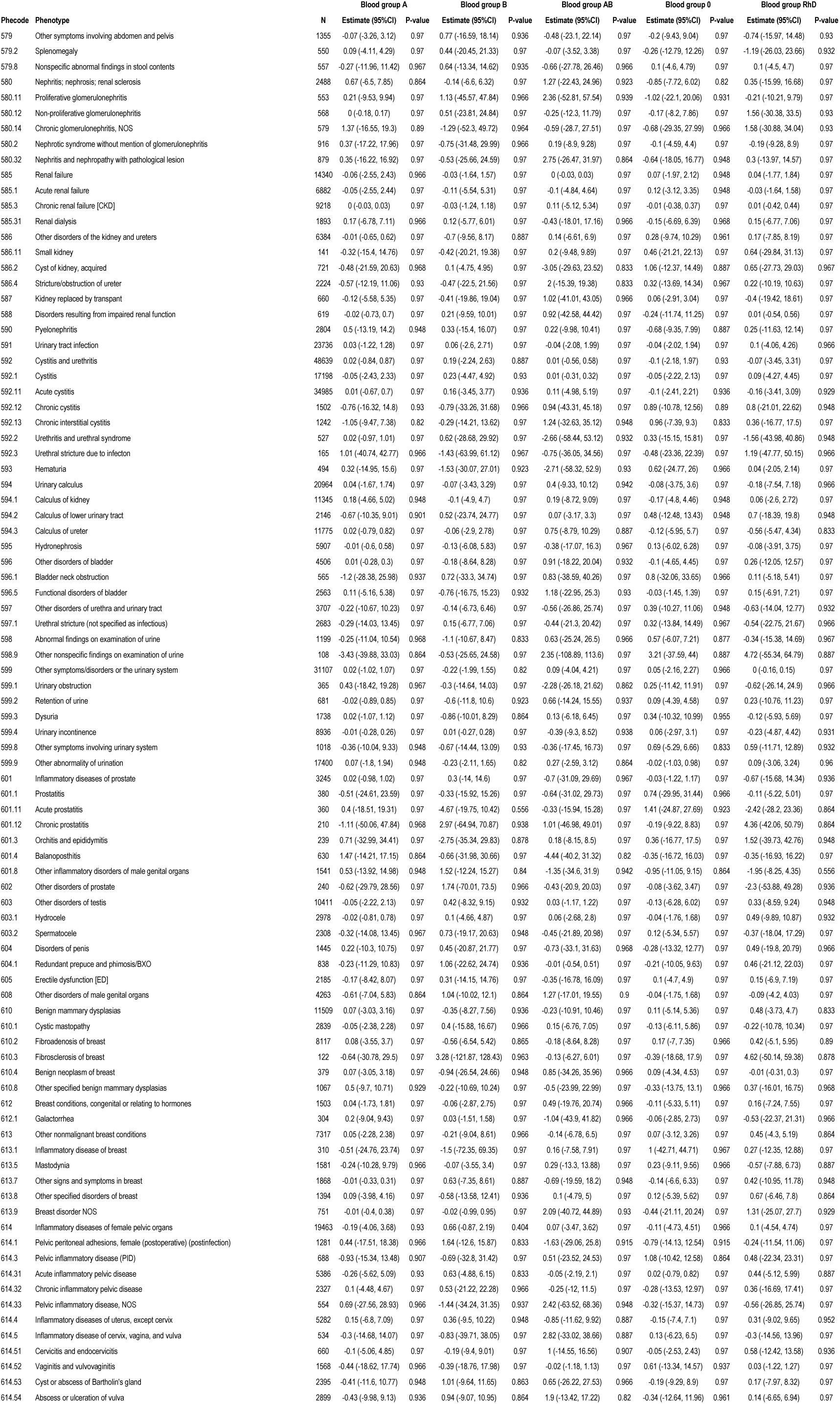
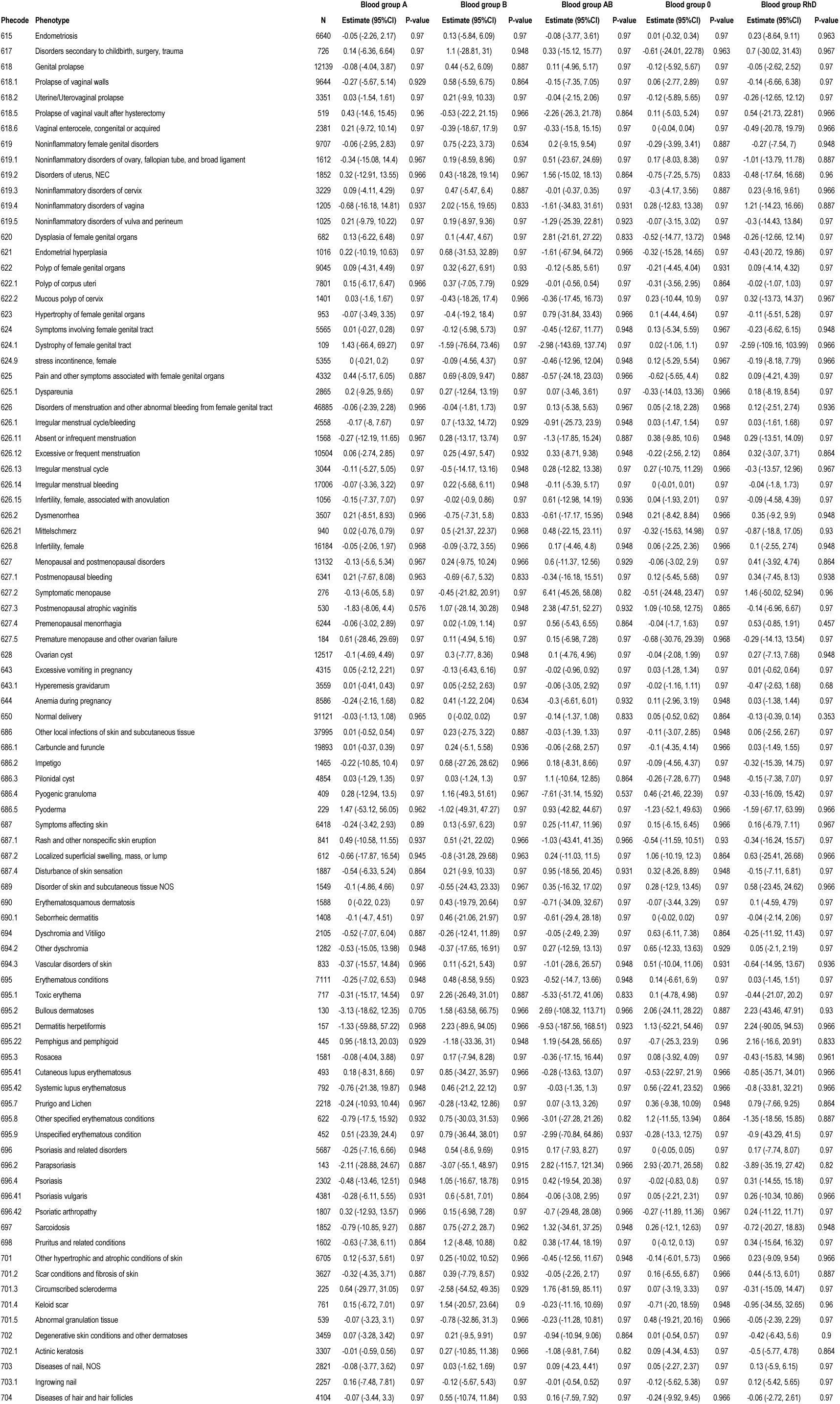
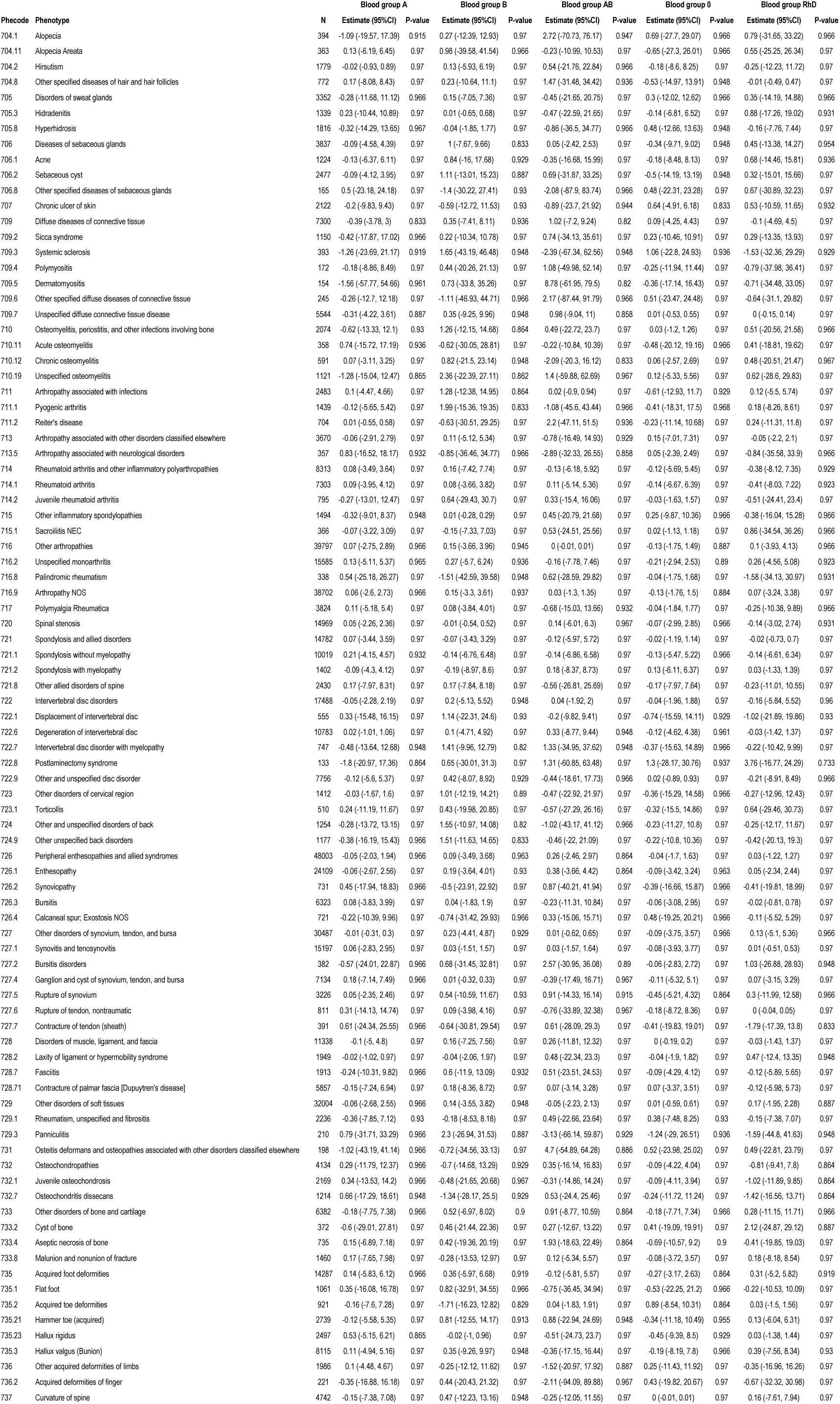
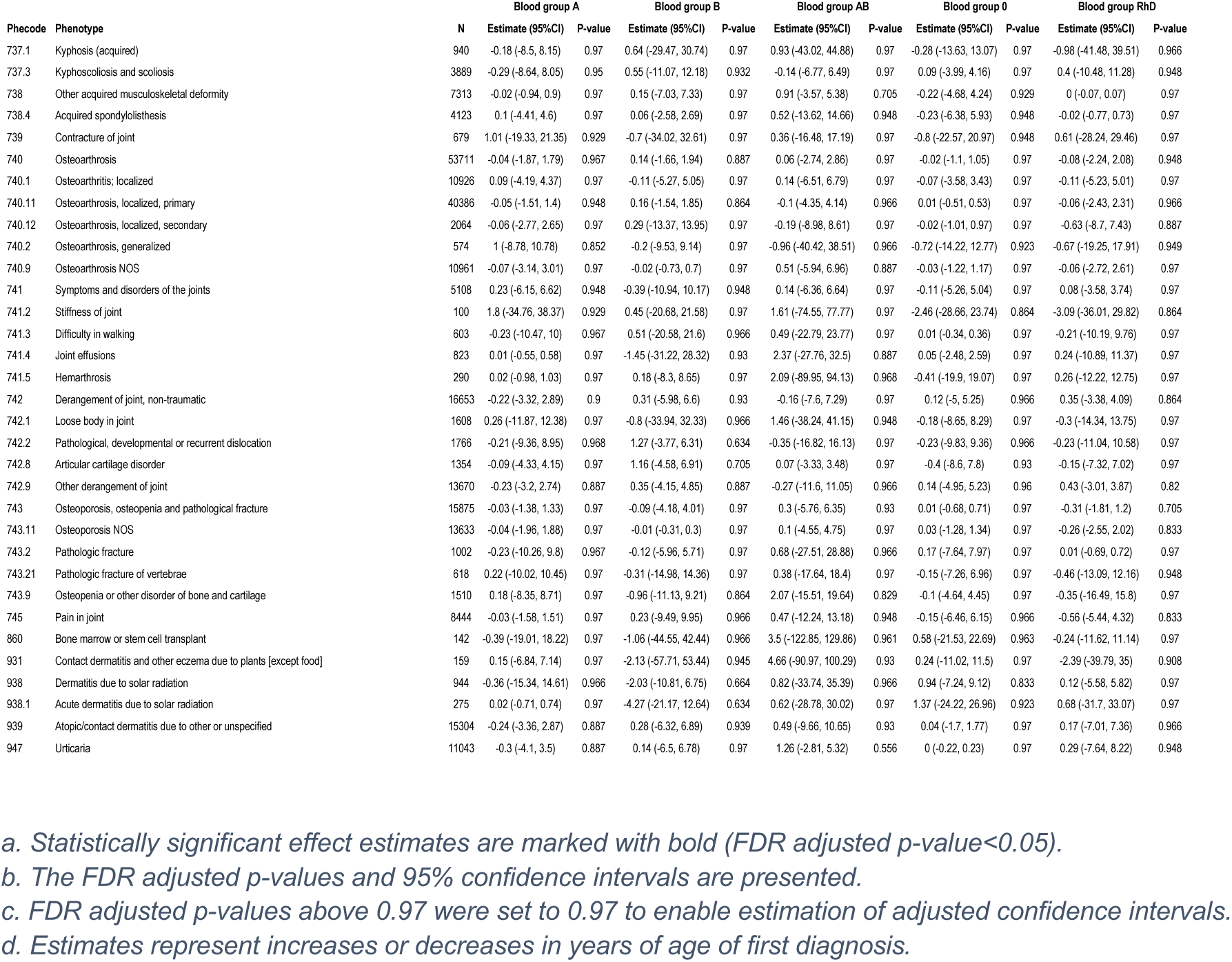
Associations between ABO/RhD blood groups and the age of the first diagnosis

